# Diffusion tensor imaging in Chediak Higashi Disease

**DOI:** 10.1101/2025.08.06.25332949

**Authors:** Connor J. Lewis, Selby I. Chipman, Cynthia J. Tifft, Camilo Toro, William A. Gahl, Maria T. Acosta, Wendy J. Introne

## Abstract

**AIM:** To define the natural history of diffusion tensor imaging (DTI) in Chediak-Higashi Disease (CHD) participants in relation to normative development.

**Methods:** Twenty-five DTI scans from 15 CHD participants were compared with 100 DTI scans from 100 neurotypical controls (NC). Comparisons were evaluated for DTI metrics including fractional anisotropy (FA), and mean, axial, and radial diffusivity (MD, AD, RD, respectively). Correlational tractography was also performed to identify group differences between CHD and NC.

**Results:** Pediatric CHD participants had DTI metrics similar to NC, however, progressive pathogenic increases in MD, AD, and RD were observed in CHD participants compared to NC in white matter pathways of the whole brain, corpus callosum, and cerebellum. Correlational fiber tractography identified fiber tracts with decreased FA, and increased MD, AD, and RD in CHD compared to NC.

**Interpretation:** Progressive deviations in DTI metrics highlight the progressive neurodegeneration of CHD. Aberrations in cerebellar white matter is reflective of the clinical neurologic phenotype of CHD including cerebellar dysfunction and cognitive decline. The corollary of progressive aberrations in DTI metrics and progressive clinical neurodegeneration suggest DTI may be a suitable neuroimaging marker for CHD. Future studies should evaluate functional magnetic resonance imaging and volumetric studies in this cohort.

## Introduction

Chediak-Higashi Disease (CHD) is an ultra-rare genetic disease characterized by oculocutaneous albinism, bleeding diathesis, congenital immunodeficiency, hemophagocytic lymphohistiocytosis (HLH), neurodevelopmental deficits and neurodegeneration after the first decade of life.^1^ CHD is caused by biallelic variants in the lysosomal trafficking regulator gene, *LYST*.^1^ CHD was first described more than eight decades ago, but the function of *LYST* remains largely a mystery. CHD is extremely rare with fewer than 500 cases reported worldwide.^1^

Bone marrow transplantation corrects the hematologic and immune system deficiencies of CHD and protects against HLH, also called the accelerated phase.^2^ If untreated, these complications lead to high mortality in the first decade of life.^2^ Bone marrow transplantation, however, does not address the progressive CNS tissue damage that causes neurodevelopmental difficulties and later cerebellar dysfunction, peripheral sensorimotor neuropathy, parkinsonism, cognitive decline and premature death.^3,4,5,6^

MRI data are limited in CHD participants, but often reflect the acute inflammation and demyelination associated with complications of HLH.^7,8^ One case series, however, described MRI findings in 9 individuals with CHD who never had HLH.^9^ Cerebral and/or cerebellar atrophy were the most frequent findings and appeared more severe in participants with advanced CHD.^9^ Other findings include diminished volume of the posterior fossa, suggesting cerebellar hypoplasia, and features of cerebellar atrophy.^9^ These findings might correspond to the neurodevelopmental and neurodegenerative phases experienced by CHD participants. To our knowledge, no previous studies have quantified diffusion tensor imaging (DTI) metrics in CHD.

DTI is a neuroimaging technique utilized in neurosurgical planning, tumor cell identification, stroke progression, and neurodegenerative diseases.^10,11^ Using multiple radio frequency pulses, water’s diffusion can be characterized *in vivo* to quantify DTI metrics including fractional anisotropy (FA), mean diffusivity (MD), axial diffusivity (AD), and radial diffusivity (RD). Increased MD and decreased FA are associated with damage or disruption in fiber integrity,^12^ while increased RD and AD are associated with a loss of myelin integrity and reduced axonal density, respectively.^13^

Correlational fiber tractography, a group-level DTI analysis, can identify correlations between a study variable and variations in a DTI metric.^14^ We adapted this technique to identify differences between cohorts; in a previous study, we characterized the cerebellar pathology in adults with Tay-Sachs and Sandhoff disease.^15^ In the current study, we employed DTI to describe the natural history of the neuroimaging CHD phenotype compared to neurotypical development.

## Methods

### CHD Participants

Participants with a confirmed diagnosis of CHD and at least one DTI scan enrolled in NCT00005917,^16^ approved by the NIH Institutional Review Board (00-HG-0153) were included. All patient visits were conducted at the NIH Clinical Center between 2011 and 2024. Twenty-five DTI scans from 15 CHD participants were included. 4 CHD participants were less than 5 years old, and the remaining participants were adults.

### Neurotypical Controls

Four age- and sex-matched neurotypical controls were selected for each CHD DTI scan totaling 100 participants for the DTI NC cohort (Appendix A for details).

### DTI Preprocessing

DTI acquisition parameters are provided in Appendix B. As described in Lewis et al.^15^ Unprocessed DTI were preprocessed using MRtrix3’s (MRtrix, v3.0.4) *dwifslpreproc* command.^17^ DSI Studio (DSI Studio, v2023) was used to perform q-space diffeomorphic reconstruction with a diffusion sampling length ratio of 1.25.^15,17,18^

### DTI Analysis

The 125 preprocessed and reconstructed DTI scans were analyzed with a diffusion MRI connectometry to calculate diffusion tensor metrics. An atlas based deterministic fiber tractography was performed for the whole brain, cerebellum (bilateral), inferior cerebellar peduncle (bilateral), middle cerebellar peduncle (MCP), superior cerebellar peduncle (SCP), vermis, and corpus callosum to calculate FA, MD, AD, and RD. The angular threshold was 0 (random), the step size was 0 (random) mm. Tracks < 20 mm or > 200 mm were discarded, and 1,000,000 seeds were placed.

### Correlational Tractography Analysis

A separate diffusion MRI connectometry analysis was performed in DSI Studio (DSI Studio, v2023) to identify differences in DTI parameters between CHD participants and NC. Correlational tractography was performed comparing DTI scans from CHD participants and NC. A *T*-score between 2.0-4.0 was assigned for deterministic fiber tractography for the correlation between cohort designation and each of the DTI parameters. A length threshold between 10-30 voxels was also applied and fiber tracts shorter than the threshold were removed. The effects of age and sex were removed using a multiple regression model, and the false detection rate (FDR) was estimated using 4,000 random permutations and a threshold of <0.05.^15^

### Statistical Analysis

Statistical analysis was performed in R (The R Foundation, v4.3.1). Linear mixed effects models (LMEM) were built to evaluate the interaction between the presence of CHD and age on DTI derived metrics.^19,20,21^ The effect of sex was modeled as a covariate of no interest where males were assigned a value of 1 and females were assigned a value of 0. A subject level random intercept was used to account for repeated DTI scans conducted for each CHD participant. CHD participants were assigned a value of 1 and NC controls were assigned a value of 0 for cohort designation to test the effect of CHD on neurotypical development. The interaction between age and the cohort comparison variable was evaluated using a likelihood-ratio test.^20,21^ Sample LMEM code is included in Appendix C. *P*-values < 0.05 were considered significant after a Bonferroni correction for multiple comparisons.

## Results

### 3.1 DTI Analysis of FA

Table 1. summarizes the DTI results of FA between CHD and NC participants as evaluated by LMEM. There was no statistically significant interactive effect between participant age and the presence of CHD on whole brain (*χ*^2^(1) = 7.12, *p*-value_corrected_ = 0.2736, Figure 1A), corpus callosum (*χ*^2^(1) = 7.96, *p*-value_corrected_ = 0.1728, Figure 2A), or superior cerebellar peduncle (*χ*^2^(1) = 0.01, *p*-value_corrected_ = 1.000, Figure 3A) FA. There was no effect of the interactions of participant age and the presence of CHD on FA in the remaining six cerebellar pathways (Table 1, Appendix D).

**Figure 1.**
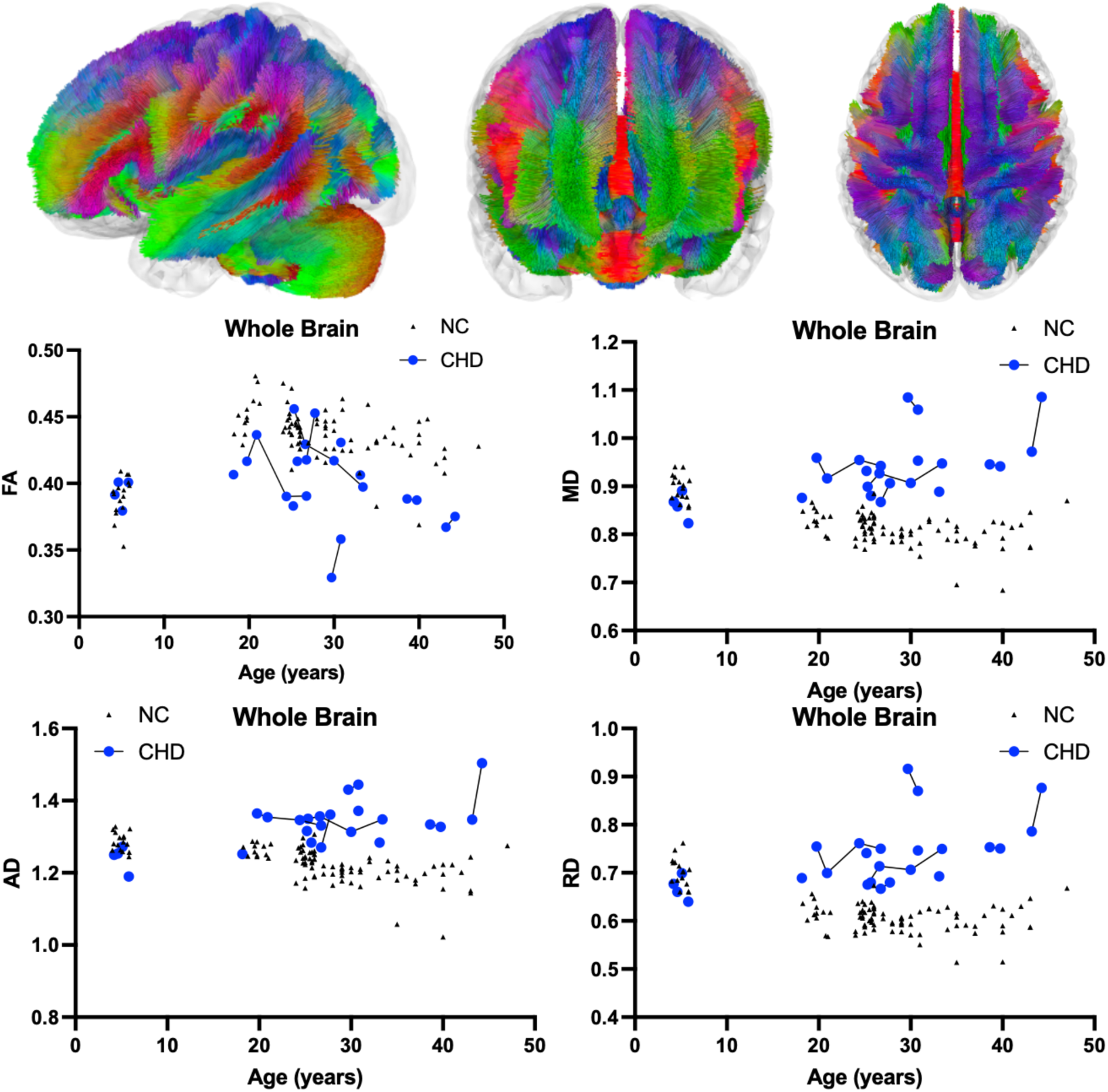
Atlas Based Fiber Tractography of the whole brain demonstrating age related effects on A.) fractional anisotropy (FA), B.) mean diffusivity (MD), C.) radial diffusivity (RD), D.) axial diffusivity (AD) between CHD participants (blue) and NC controls (black triangles). There was no statistically significant interaction between participant age and the presence of CHD on whole brain FA (*χ*^2^(1) = 7.12, *p*-value_corrected_ = 0.2736). There was a statistically significant positive interaction between participant age and the presence of CHD on whole brain MD (*χ*^2^(1) = 58.19, *p*-value_corrected_ < 0.0001), AD (*χ*^2^(1) = 52.93, *p*-value_corrected_ < 0.0001), and RD (*χ*^2^(1) = 52.52, *p*-value_corrected_ < 0.0001).

**Figure 2.**
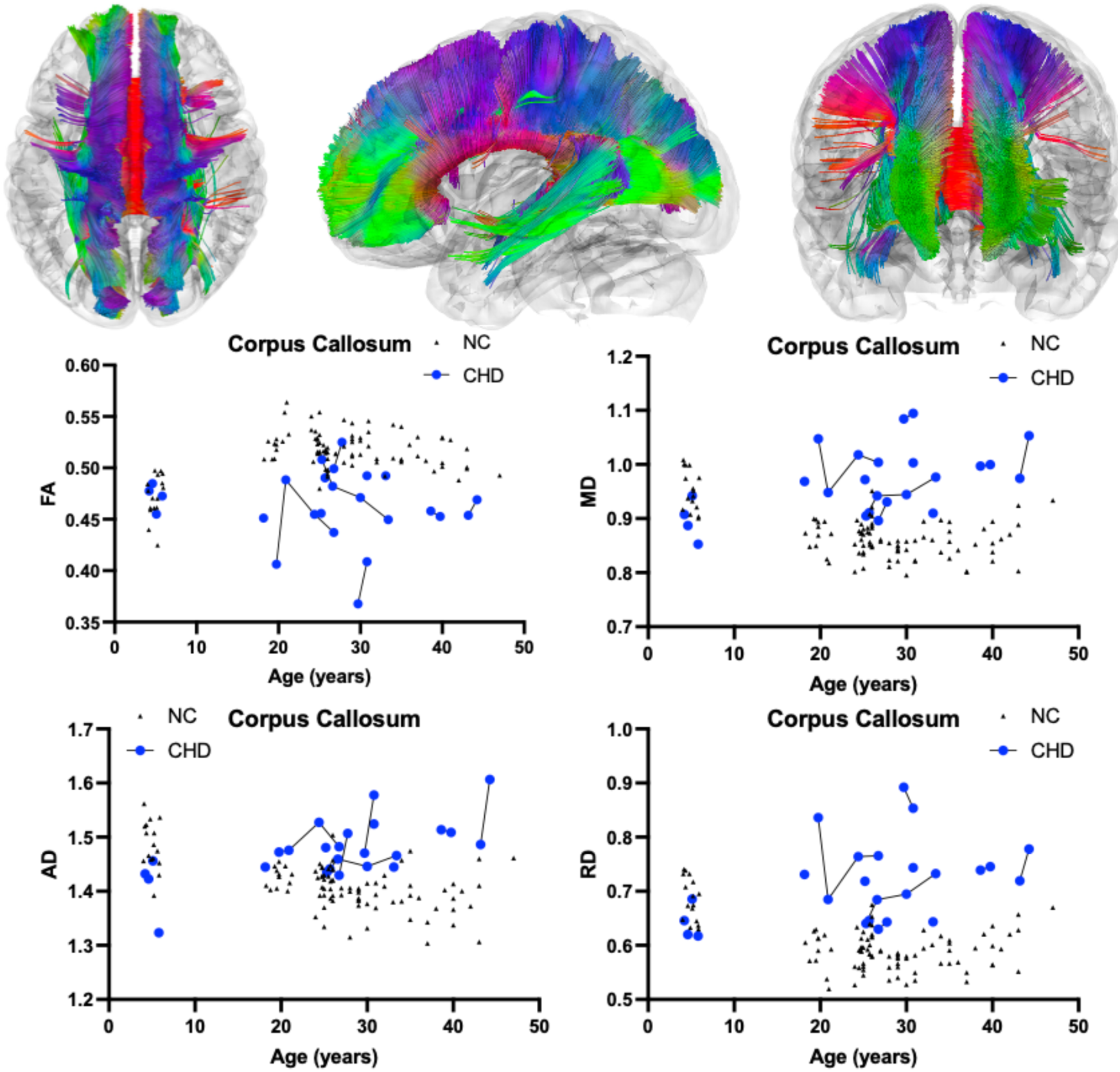
Atlas Based Fiber Tractography of the corpus callosum demonstrating age related effects on A.) fractional anisotropy (FA), B.) mean diffusivity (MD), C.) radial diffusivity (RD), D.) axial diffusivity (AD) between CHD participants (blue) and NC controls (black triangles). There was no statistically significant interaction between participant age and the presence of CHD on corpus callosum FA (*χ*^2^(1) = 7.96, *p*-value_corrected_ = 0.1728). There was a statistically significant negative interaction between participant age and the presence of CHD on corpus callosum MD (*χ*^2^(1) = 37.67, *p*-value_corrected_ < 0.0001), AD (*χ*^2^(1) = 46.87, *p*-value_corrected_ < 0.0001), and RD (*χ*^2^(1) = 27.52, *p*-value_corrected_ < 0.0001).

**Figure 3.**
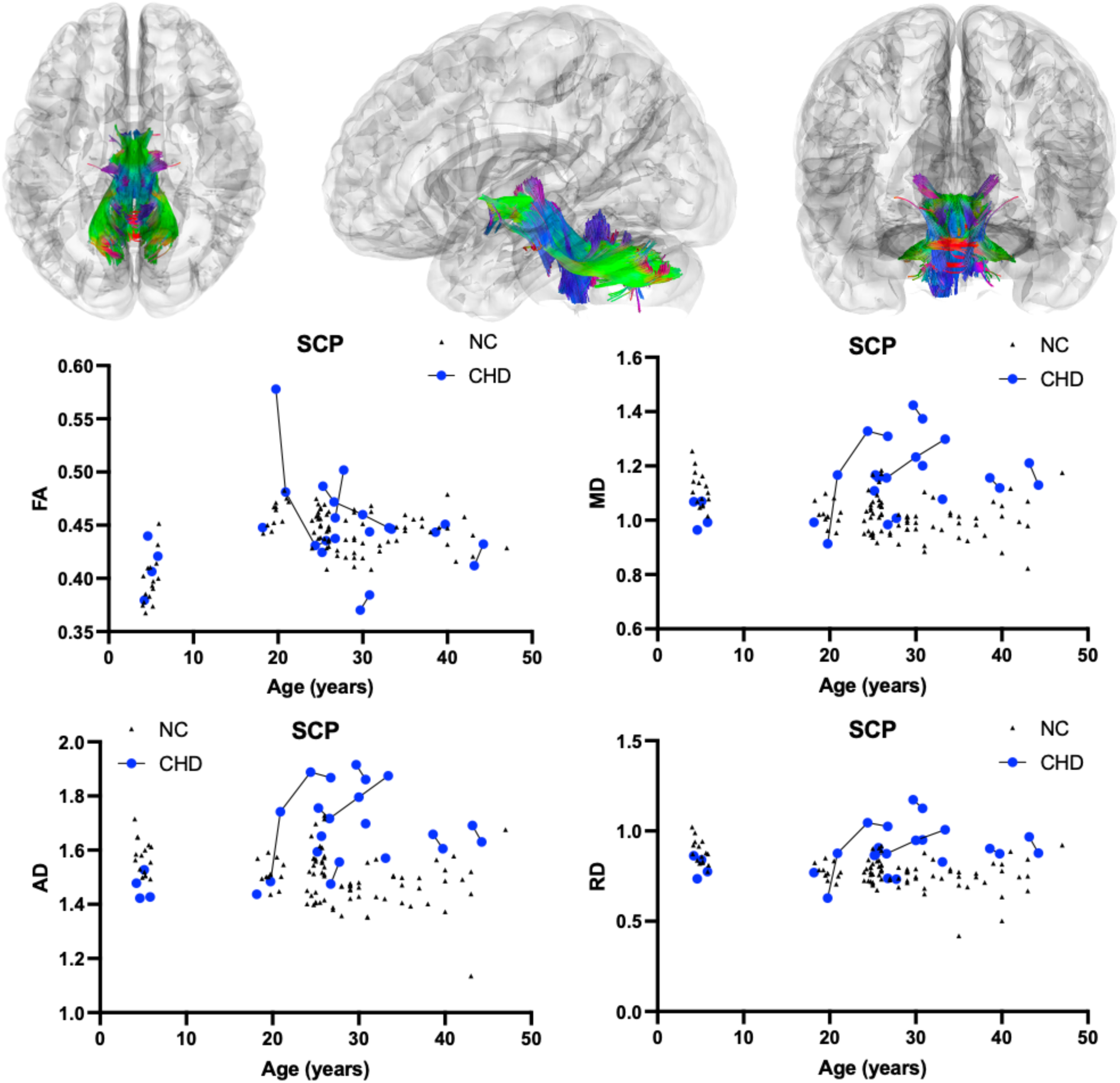
Atlas Based Fiber Tractography of the superior cerebellar peduncle (SCP) demonstrating age related effects on A.) fractional anisotropy (FA), B.) mean diffusivity (MD), C.) radial diffusivity (RD), D.) axial diffusivity (AD) between CHD participants (blue) and NC controls (black triangles). There was no statistical effect of the interaction between participant age and the presence of CHD on SCP FA (*χ*^2^(1) = 0.01, *p*-value_corrected_ = 1.000). There was a statistically significant negative interaction between participant age and the presence of CHD on SCP MD (*χ*^2^(1) = 21.36, *p*-value_corrected_ = 0.0001), AD (*χ*^2^(1) = 17.75, *p*-value_corrected_ = 0.0009), and RD (*χ*^2^(1) = 21.73, *p*-value_corrected_ = 0.0001).

**Table 1.** Diffusion Tensor Imaging results in fiber tractography pathways evaluating differences between NC and CHD participants. Estimates and Standard Errors for all model variables are included in Section E of the Supplementary Appendix. FA= Fractional Anisotropy; MD= Mean Diffusivity, AD = Axial Diffusivity, RD = Radial Diffusivity.

| Pathway | Estimate | Standard Error | $\chi^2(1)$ | $p$-value ($>\chi^2$) | $p$-value (corrected) |
| --- | --- | --- | --- | --- | --- |
| FA |  |  |  |  |  |
| Whole Brain | -0.0014 | 0.0005 | 7.12 | 0.0076 | 0.2736 |
| Left Cerebellum | -0.0006 | 0.0008 | 0.47 | 0.4908 | 1.000 |
| Right Cerebellum | < 0.0001 | 8.7e-04 | < 0.01 | 0.9864 | 1.000 |
| Left Inferior Cerebellar Peduncle | -0.0012 | 0.0013 | 0.79 | 0.3746 | 1.000 |
| Right Inferior Cerebellar Peduncle | -0.0001 | 0.0014 | 0.30 | 0.5822 | 1.000 |
| Middle Cerebellar Peduncle | -0.0009 | 0.0015 | 0.31 | 0.5794 | 1.000 |
| Superior Cerebellar Peduncle | -0.0001 | 0.0011 | 0.01 | 0.9380 | 1.000 |
| Vermis | 0.0016 | 0.0014 | 1.46 | 0.2265 | 1.000 |
| Corpus Callosum | -0.0014 | 0.0005 | 7.96 | 0.0048 | 0.1728 |
| MD |  |  |  |  |  |
| Whole Brain | 0.0065 | 0.0008 | 58.19 | < 0.0001 | < **0.0001** |
| Left Cerebellum | 0.0113 | 0.0024 | 21.73 | < 0.0001 | **0.0001** |
| Right Cerebellum | 0.0105 | 0.0024 | 18.59 | < 0.0001 | **0.0006** |
| Left Inferior Cerebellar Peduncle | 0.0094 | 0.0026 | 13.06 | 0.0003 | **0.0109** |
| Right Inferior Cerebellar Peduncle | 0.0091 | 0.0026 | 12.39 | 0.0004 | **0.0155** |
| Middle Cerebellar Peduncle | 0.0114 | 0.0025 | 20.23 | < 0.0001 | **0.0002** |
| Superior Cerebellar Peduncle | 0.0101 | 0.0021 | 21.36 | < 0.0001 | **0.0001** |
| Vermis | 0.0141 | 0.0039 | 13.16 | 0.0003 | **0.0103** |
| Corpus Callosum | 0.0056 | 0.0009 | 37.67 | < 0.0001 | < **0.0001** |
| AD |  |  |  |  |  |
| Whole Brain | 0.0070 | 0.0009 | 52.93 | < 0.0001 | < **0.0001** |
| Left Cerebellum | 0.0119 | 0.0029 | 15.51 | < 0.0001 | **0.0030** |
| Right Cerebellum | 0.0118 | 0.0031 | 13.31 | 0.0003 | **0.0095** |
| Left Inferior Cerebellar Peduncle | 0.0089 | 0.0034 | 6.672 | 0.0098 | 0.3537 |
| Right Inferior Cerebellar Peduncle | 0.0084 | 0.0035 | 5.723 | 0.0168 | 0.6030 |
| Middle Cerebellar Peduncle | 0.0114 | 0.0033 | 11.66 | 0.0006 | **0.0230** |
| Superior Cerebellar Peduncle | 0.0132 | 0.0031 | 17.75 | < 0.0001 | **0.0009** |
| Vermis | 0.0147 | 0.0048 | 8.783 | 0.0030 | 0.1090 |
| Corpus Callosum | 0.0060 | 0.0008 | 46.87 | < 0.0001 | < **0.0001** |
| RD |  |  |  |  |  |
| Whole Brain | 0.0064 | 0.0008 | 52.52 | < 0.0001 | < **0.0001** |
| Left Cerebellum | 0.0116 | 0.0021 | 27.04 | < 0.0001 | < **0.0001** |
| Right Cerebellum | 0.0115 | 0.0021 | 25.79 | < 0.0001 | < **0.0001** |
| Left Inferior Cerebellar Peduncle | 0.0094 | 0.0022 | 16.84 | < 0.0001 | **0.0015** |
| Right Inferior Cerebellar Peduncle | 0.0089 | 0.0022 | 15.65 | < 0.0001 | **0.0027** |
| Middle Cerebellar Peduncle | 0.0105 | 0.0020 | 24.89 | < 0.0001 | < **0.0001** |
| Superior Cerebellar Peduncle | 0.0086 | 0.0018 | 21.73 | < 0.0001 | **0.0001** |
| Vermis | 0.0136 | 0.0036 | 14.02 | 0.0002 | **0.0072** |
| Corpus Callosum | 0.0054 | 0.0010 | 27.52 | < 0.0001 | < **0.0001** |

### 3.2 DTI Analysis of MD

As described in Table 1, there was a statistically significant positive interactive effect between age and presence of CHD on whole brain (*χ*^2^(1) = 58.19, *p*-value_corrected_ < 0.0001, Figure 1B), corpus callosum (*χ*^2^(1) = 46.87, *p*-value_corrected_ < 0.0001, Figure 2B), and superior cerebellar peduncle (*χ*^2^(1) = 21.36, *p*-value_corrected_ = 0.0001, Figure 3B) MD. There was also a significant positive interactive effect between age and the presence of CHD in the remaining analyzed cerebellar pathways including the bilateral cerebellum, bilateral inferior cerebellar peduncle, middle cerebellar peduncle, and vermis (Table 1, Appendix D).

### 3.3 DTI Analysis of AD

Table 1 also shows a statistically significant positive interactive effect between age and presence of CHD on whole brain (*χ*^2^(1) = 52.93, *p*-value_corrected_ < 0.0001, Figure 1C), corpus callosum (*χ*^2^(1) = 46.87, *p*-value_corrected_ < 0.0001, Figure 2C), and superior cerebellar peduncle (*χ*^2^(1) = 21.36, *p*-value_corrected_ = 0.0009, Figure 3C) AD. There was a significant positive interactive effect between age and the presence of CHD in three of the six remaining analyzed cerebellar pathways including the bilateral cerebellum and middle cerebellar peduncle (Table 1, Appendix D).

### 3.4 DTI Analysis of RD

As described in Table 1, there was a statistically significant positive interactive effect between age and presence of CHD on whole brain (*χ*^2^(1) = 52.52, *p*-value_corrected_ < 0.0001, Figure 1D), corpus callosum (*χ*^2^(1) = 46.87, *p*-value_corrected_ < 0.0001, Figure 2D), and superior cerebellar peduncle (*χ*^2^(1) = 21.73, *p*-value_corrected_ = 0.0001, Figure 3D) RD. There was also significant positive interactive effect between age and the presence of CHD in the remaining analyzed cerebellar pathways including the bilateral cerebellum, bilateral inferior cerebellar peduncle, middle cerebellar peduncle, and vermis (Table 1, Appendix D).

### 3.5 Correlational Fiber Tractography Analysis

Figure 4 shows significant differences in FA in fiber tracts between CHD and NC evaluated by correlational fiber tractography. At low *T*-score and length thresholds, NC participants exhibited numerous fiber tracts (112,845 tracts) with higher FA compared to CHD participants. Fiber tracts with higher FA in NC were identified primarily in association pathways (54 percent of tracts with higher FA in NC) and commissure (corpus callosum, 32 percent) pathways (Supplement Table H1). Specifically, regions with the highest percentage of fiber tracts were the bilateral inferior fronto-occipital fasciculus (19.15 percent), corpus callous tapetum (9.97 percent), and corpus callosum body (9.68 percent). At higher *T*-scores and length thresholds, considerably fewer fiber tracts (2916 tracts) were identified with higher FA in NC compared to CHD participants, primarily in commissure pathways (92.22 percent, Supplement Table H2). At low length thresholds and *T*-scores, CHD participants were evaluated to have fiber tracts (4,932 tracts) with higher FA (Figure 4), solely in cerebellar pathways. However, at higher length thresholds and *T*-scores, no fiber tracts had higher FA in CHD compared to NC.

**Figure 4.**
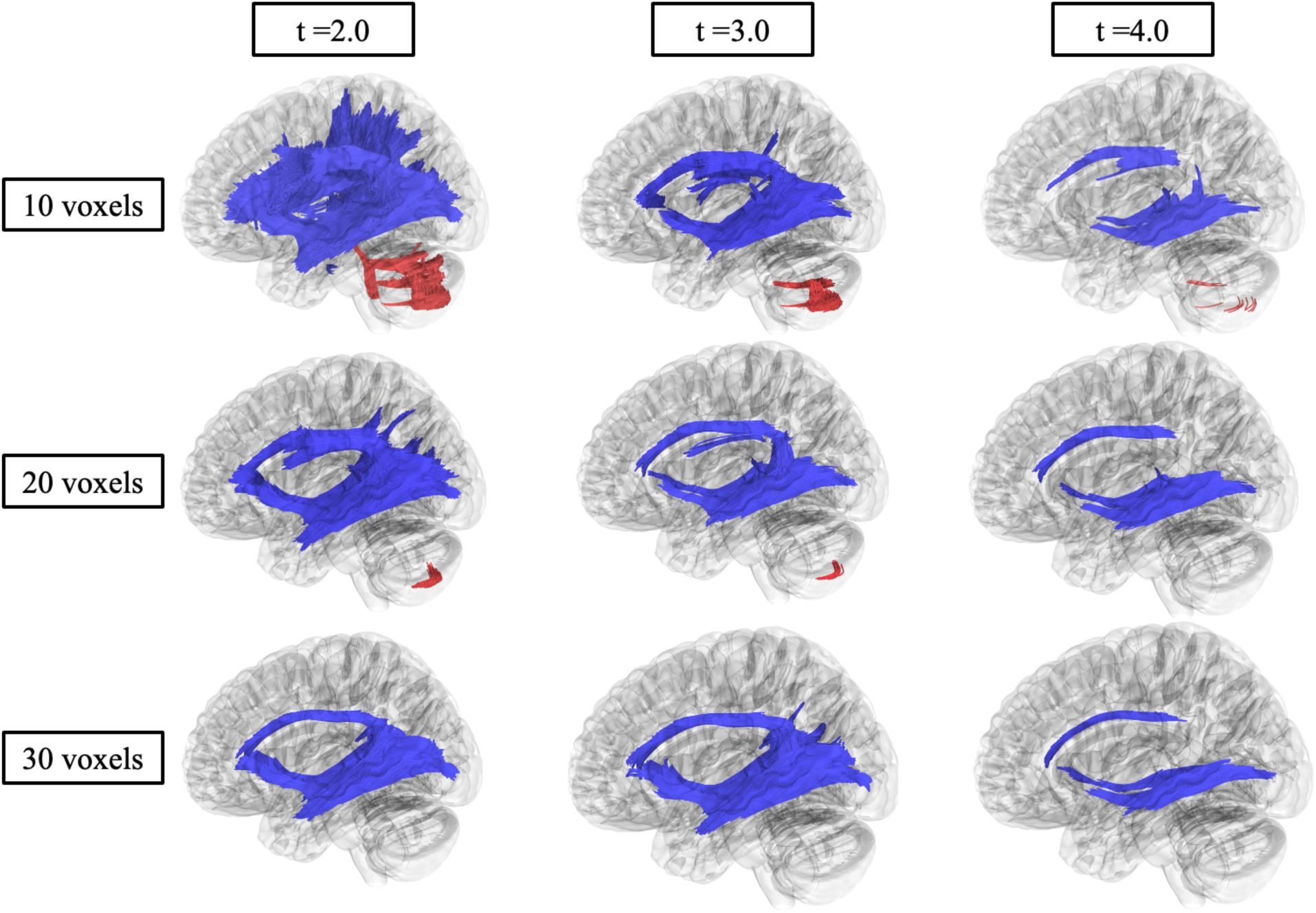
Correlational fiber tractography assessed differences in fractional anisotropy (FA) in CHD participants and NC at varying lengths (voxels) and T thresholds. Fiber tracts shown in red were evaluated to have a higher fractional anisotropy in CHD participants compared to NC participants and were observed primarily in the cerebellum (FDR < 0.05). Fiber tracts shown in blue were evaluated to have a higher FA in NC compared to CHD participants and were primarily observed in association and corpus callosum pathways.

Figure 5 show significant differences in MD in fiber tracts compared between CHD and NC as evaluated by correlational fiber tractography. Numerous fiber tracts (409,476) were identified with higher MD in CHD participants compared to NC. At low thresholds, fiber tracts were identified throughout the brain including association (33.18 percent of tracts with higher MD in CHD), commissure (19.50 percent), basal ganglia (17.28 percent), brainstem (15.68 percent), and cerebellar (14.01 percent) pathways. Specifically, tracts with the highest prevalence were the body of the corpus callosum (9.77 percent), left arcuate fasciculus (6.78 percent) tapetum of the corpus callosum (5.71 percent), and the left (5.19 percent) and right cerebellum (5.18 percent). At higher length thresholds and *T*-scores, notably fewer fiber tracts (31,029) were identified, primarily association pathways (92.79 percent). No fiber tracts were identified to have lower MD in CHD participants compared to NC.

**Figure 5.**
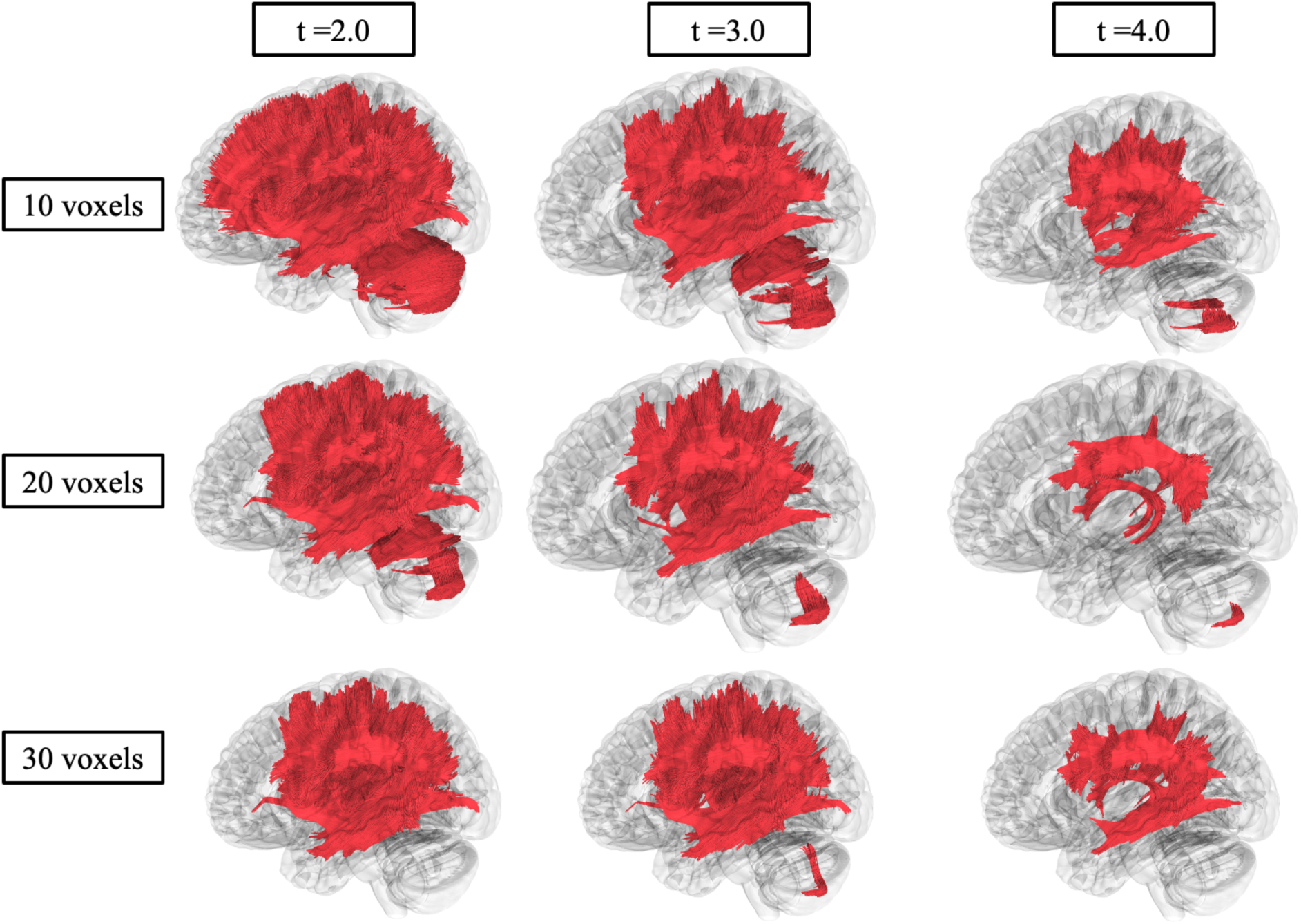
Correlational fiber tractography assessed differences in mean diffusivity (MD) in CHD participants and NC at varying length (voxels) and T threshold. Fiber tracts shown in red were evaluated to have a higher mean diffusivity in CHD participants compared to NC participants and were observed primarily in the corpus callosum, association, and cerebellar pathways (FDR < 0.05). No fiber tracts were evaluated to have a higher MD in NC compared to CHD participants.

Supplementary Figure G1 shows significant differences in AD between CHD and NC evaluated by correlational fiber tractography. At low *T*-score and length thresholds, numerous fiber tracts (134,693 tracts) had higher AD in CHD participants compared to NC primarily in cerebellar (48.29 percent of tracts with higher AD in CHD), basal ganglia (18.61 percent), and brainstem (17.56 percent) pathways. The pathways with the highest prevalence of tracts were the left (19.52 percent) and right cerebellum (17.79 percent), the middle cerebellar peduncle (5.55 percent), and the tapetum corpus callosum (4.39 percent). At high *T*-score and length thresholds, notably fewer fiber tracts had higher AD in CHD participants compared to NC (87 total tracts) and were identified solely in basal ganglia projection pathways. No fiber tracts had lower AD in CHD participants compared to NC.

Similar results to MD were observed for RD (Supplement Figure G2) as evaluated by correlational tractography. Numerous fiber tracts (382,315 tracts) with higher RD were identified in CHD participants compared to NC at low *T*-scores and length thresholds. These fiber tracts were spread across the brain including the association (42.89 percent), commissure (24.42 percent), basal ganglia (15.31 percent), cerebellar (8.81 percent), and brainstem (8.55 percent) pathways. The pathways with the highest prevalence of tracts were the body (11.34 percent) and tapetum (6.77 percent) of the corpus callosum, left arcuate fasciculus (6.52 percent), and right inferior fronto-occipital fasciculus (5.96 percent). At higher *T*-scores and length thresholds, fewer fiber tracts (30,865 tracts) were identified to have higher RD in CHD compared to NC. These tracts were identified primarily in association (82.90 percent) and basal ganglia (10.31 percent) pathways. No fiber tracts were identified to have lower RD in CHD participants compared to NC.

## Discussion

In this study, we aimed to define the natural history of DTI in CHD participants in relation to neurotypical development. The statistically significant interactions between age and presence of CHD on DTI metrics demonstrated progressive neurodegeneration of CHD. Pediatric CHD participants showed lower MD, AD, and RD values similar to their NC, whereas adult CHD participants show pathogenic increases in these DTI metrics. Notably, increased MD, AD, and RD were observed throughout whole brain and corpus callosum fiber tracts (Figures 1 and 2). Although no other study has reported DTI derived metrics in CHD participants, this result is consistent with previous studies noting atrophy of the cerebrum and corpus callosum.^22,23^ These DTI results are consistent with the recognized clinical and neurologic phenotypes of CHD.

Shirazi et al.^24^ reported no cognitive impairments in pediatric participants with CHD who had received bone marrow transplantation; however, adults with CHD exhibited significant cognitive impairments across all domains tested.^24^ Their study, however, reported results from only 4 CHD participants less than 5 years old, and many of their adult participants had ADHD and/or required an individualized educational plan in childhood.^24^

Furthermore, there were progressive abnormal increases in MD, AD, and RD throughout pathways of the cerebellum including the bilateral cerebellum, bilateral inferior cerebellar peduncle, middle cerebellar peduncle, superior cerebellar peduncle, and vermis. The cerebellum has been implicated in mouse models of CHD showing Purkinje cell damage and causing cerebellar neurodegeneration and deficits.^25,26^ Clinical cerebellar deficits documented in CHD include cerebellar ataxia, balance difficulties, action tremors, nystagmus, and dysmetria,^9,27^ suggesting that cerebellar DTI metrics reflect the cerebellar degeneration in CHD. Disruptions of the cerebellar output (superior cerebellar peduncle) to the thalamus as a part of the cortico-cerebellar-cortical networks may contribute to the cognitive, behavioral, and motor symptoms of CHD participants. Additionally, DTI results show that progression of CHD may resemble Parkinson’s disease’s effects on the same DTI metrics in the brain, which is noteworthy since CHD can cause L-Dopa responsive parkinsonism in some participants.^5,27,28^ Ultimately, the DTI metrics evaluated in this study parallel the clinical neurologic phenotype of CHD participants, suggesting that DTI is a suitable neuroimaging surrogate marker for potential therapeutic interventions.

While there were no statistically significant differences in FA between CHD participants and NC after the Bonferroni correction for multiple comparisons, correlational fiber tractography did show numerous significant differences in FA between CHD and NC. Decreased FA was observed in CHD participants in association pathways including the bilateral inferior fronto-occipital fasciculus and corpus callosum. The inferior fronto-occipital fasciculus, a bundle of fiber tracts connecting the occipital and parietal lobes to the frontal lobe, affect language processing, goal-oriented behavior, and visual switching tasks.^29^ Degeneration of the inferior fronto-occipital fasciculus has been implicated in behavioral and neurological disorders.^29,30,31^ Regarding the present study, alterations in the microstructure of the corpus callosum may be related to the motor and cognitive impairments experienced by CHD participants.

Correlational tractography of MD, AD, and RD all showed significant differences between CHD and NC. Deviations in MD were present in the corpus callosum, arcuate fasciculus, and cerebellum, which support the DTI results of the cerebellum and corpus callosum. The arcuate fasciculi and corpus callosum are involved in intra- and trans-cortical connectivity as a part of the association pathway and commissure pathways, respectively. Dysfunctions in these pathways likely disrupt higher cortical function in CHD participants.

Disruptions of multiple pathways were found in CHD compared to NC, which could explain cognitive impairments in this group. While alterations in a single pathway could not explain the full neuropsychological phenotype of CHD, a more plausible explanation is the disruption of multiple functional intracortical, transcortical, and cortical-cerebellar-cortical networks. To our knowledge, there have been no functional MRI studies in CHD; future studies should investigate how potential impairment of functional networks, particularly in CHD adults, relate to both the neurological and neuropsychological phenotype of CHD alongside structural aberrations.

Limitations of this study should be considered before using the results as outcome measures in clinical trials or in clinical practice. First, the study involved a small sample size with 15 CHD participants being included in the DTI analysis. However, CHD is an ultra-rare disorder with fewer than 500 cases reported,^1^ so this analysis incorporated 3% of reported cases.

The study involves variable DTI acquisition protocols. CHD participants were scanned with 15 diffusion directions compared to 24-64 directions in NC. While a previous study showed 18 diffusion directions appears to be the minimum number to limit variations in DTI metrics, fiber tracts impacted from the 15-direction data appear limited to the corticospinal tract, superior longitudinal fasciculus, cingulate gyrus, and uncinate fasciculus, and may be less influential for MD, AD, and RD compared to FA.^32^ Pediatric NC were gathered exclusively one NC dataset, which were scanned at a lower b-value compared to CHD participants and older NC cohorts. A previous study comparing b-values found FA in the internal capsule was influenced by a low signal to noise ratio in that region.^33^ The pathways implicated by variations in the number and strength of the b-values do not appear related to the pathways indicated in this study by CHD; some caution may be warranted.

Skepticism around FA as a DTI metric have been described.^34^ For example, FA changes in both directions (or stability) can be influenced by crossing or branching fibers, leading to incorrect interpretations.^34^ We potentially observed this in both our DTI and correlational fiber tractography analysis. For instance, correlational fiber tractography investigations of FA differences between NC and CHD showed higher FA in cerebellar fiber tracts in CHD participants (Figure 4), as unexpected. This result was echoed in our DTI analysis, as there were no differences observed between CHD participants and NC in FA of cerebellar pathways (Table 1). Since MD, RD, and AD increases were observed in the same regions for both DTI and correlational tractography analyses, it is plausible that the increases in FA observed in CHD participants were due to crossing fibers and not neuronal microstructure changes. This result highlights the importance of evaluating and drawing conclusions from the combination of FA, MD, AD, and RD rather than from one DTI metric. The role of complex neuronal fiber orientation requires further investigation.

In conclusion, we documented the first cohort analysis of DTI in CHD. We found significant alterations in DTI parameters including MD, AD, and RD compared to neurotypical controls. These disruptions in white matter networks were progressive and consistent with the neurologic and neuropsychiatric phenotype of CHD. Pediatric CHD participants had DTI metrics consistent with their unaffected peers whereas adult CHD participants had significant white matter disruption. Furthermore, correlational tractography showed significant differences in FA, MD, AD, and RD between CHD participants and NC, further establishing DTI as a neuroimaging marker of disease progression in CHD. Future studies should consider detailed longitudinal volumetric data including both gray and white matter structures and functional magnetic resonance imaging to investigate discreet targets of neurodegeneration alongside potential disruptions in functional networks caused by CHD to account for the neurological deficits in this disease.

## Data Availability

Neurotypical control MRI data described in this manuscript are available from either OpenNeuro (https://openneuro.org/) or OpenscienceFramework (https://osf.io/) from the datasets mentioned in the supplement of this study. Analyzed data and data describing Chediak-Higashi Disease in this manuscript are available from the corresponding author upon reasonable request.

## Funding Statement

This work was supported by the Intramural Research Program of the National Human Genome Research Institute (Tifft ZIAHG200409). This report does not represent the official view of the National Human Genome Research Institute (NHGRI), the National Institutes of Health (NIH), the Depart of Health and Human Services (DHHS), or any part of the US Federal Government. No official support or endorsement of this article by the NHGRI or NIH is intended or should be inferred. NCT00005917.

## Acknowledgements

We thank the participants and their families from all studies for the generosity of their time, efforts, and support of open data. DTI preprocessing in this work utilized the computational resources of the Biowulf Linux cluster at the National Institutes of Health (http://hpc.nih.gov).

## Ethics Declaration

The NIH Institutional Review Board approved this protocol (00-HG-0153). Written informed consent was obtained from participants or the legal guardians of the participants. Informed consent was completed before participation, and all research was completed in accordance with the Declaration of Helsinki.

## Consent for Publication

Not applicable.

## Competing Interests

The authors declare no conflict of interest.

## Supplementary Methods

### Supplement A: Neurotypical Control Participant Characteristics

Neurotypical controls (NC) were age- and sex-matched to CHD participants (Figure B1). Four NC participants were selected for each DTI scan. 100 participants were included for the DTI analysis. Participants were selected from the following datasets:

#### Calgary Neurotypical Controls^2, 3^

16 participants (8 females) were included in the DTI analysis from the “Calgary Preschool magnetic resonance imaging (MRI)” dataset.

#### NIMH^4,5^

35 participants (12 females) were in included in the DTI analysis from “The National Institute of Mental Health (NIMH) Intramural Healthy Volunteer Dataset”

#### UCLA^6,7^

12 participants (12 males) from the “UCLA Consortium for Neuropsychiatric Phenomics LA5c Study” were included in the DTI analysis.

#### AOMIC-PIOP1^8,9^

20 participants (8 females) from the “Amsterdam Open MRI Collection (AOMIC) Population Imaging of Psychology (PIOP)” dataset were included in the DTI analysis.

#### AOMIC^8,10^

17 participants (17 males) from the “Amsterdam Open MRI Collection (AOMIC)” dataset were included in the DTI analysis.

**Figure A1.**
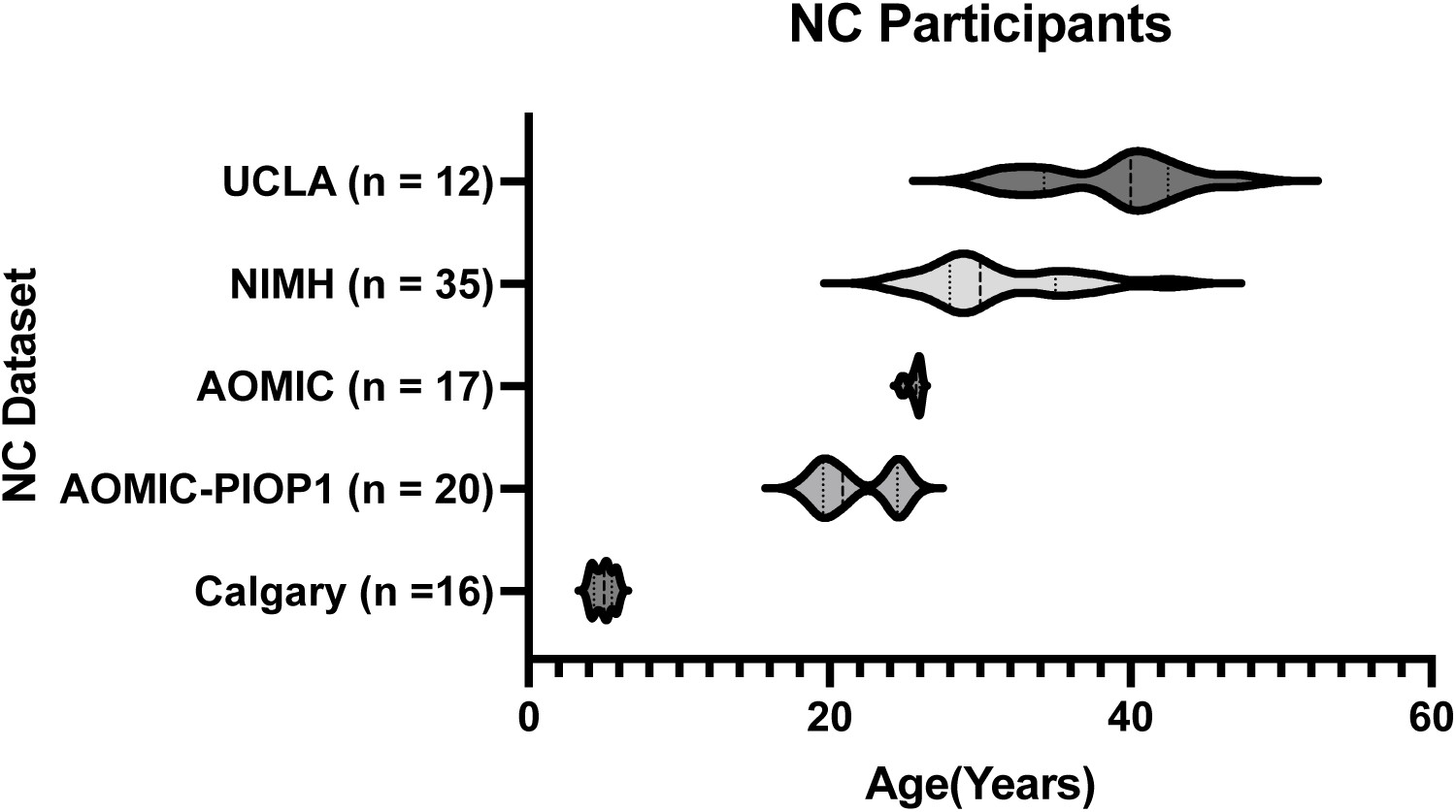
NC participant age. Each NC dataset is on a separate row for each of the five datasets used in this study.

### Supplement B: DTI Acquisition

#### Chediak-Higashi Disease Participants DTI Acquisition

DTI scans for CHD participants were conducted on a 3T Phillips (Philips Healthcare, Best, The Netherlands) Achieva MR System with an 8-channel SENSE head coil. DTI scans were collected with the following parameters: TR/TE = 6400/100 ms, 15-gradient encoding directions, b-values = 0 and 1000 s/mm^2^, voxel size = 1.875mm × 1.875mm × 2.5mm, slice thickness = 2.5 mm, acquisition matrix = 128 × 128, NEX = 1, FOV = 24 cm.

#### Calgary Neurotypical Controls^2, 3^

A General Electric 3T MR750w system with a 32-channel head coil was used for scanning all Calgary neurotypical controls using a single shot spin echo-planar imaging sequence. DTI images were acquired with the following parameters for Calgary neurotypical controls: TR/TE=6750/79 ms, 30-gradient encoding directions, b-values = 0 and 750 s/mm^2^, voxel size = 1.6mm×1.6mm×2.2mm, slice thickness = 2.2 mm, FOV = 20 cm.

#### NIMH^4,5^

A 3T General Electric 3T Discovery MR750w system with a 32-channel head coil was used for scanning all NIMH neurotypical controls using a single shot spin echo-planar imaging sequence. DTI images were acquired with the following parameters for NIMH neurotypical controls: TR/TE=7800/61 ms, 24-gradient encoding directions, b-values = 0 and 1000 s/mm^2^, voxel size = 2mm×2mm×2mm, slice thickness = 2.0 mm, FOV = 23.2 cm as modified from the Alzheimer’s Disease Neuroimaging Initiative (ADNI3).

#### UCLA^6,7^

A Siemens 3T Trio scanner was used to acquire this data. DTI using an echo-planar sequence with the following parameters for UCLA neurotypical controls: TR/TE = 9000/93 ms, 64-gradient encoding directions, b-values = 0 and 1000 s/mm^2^, voxel size = 2mm×2mm×2mm, slice thickness = 2.0 mm.

#### AOMIC-PIOP1^8,9^

A Philips 3T Achieva system with a 32-channel head coil. DTI images were acquired using single shell spin-echo diffusion-weighted imaging with the following parameters for AOMIC-PIOP1 neurotypical controls: TR/TE = 7387/86 ms, 32-gradient encoding directions, b-values = 0 and 1000 s/mm^2^, flip angle = 90°, FOV = 22.4 × 22.4 × 12.0 cm, voxel size= 2×2×2 mm, and 60 slices.

#### AOMIC^8,10^

MRI data were acquired using a Philips 3T Intera scanner with a 32-channel head coil. DTI images were acquired using single shell spin-echo diffusion-weighted imaging with the following parameters for AOMIC neurotypical controls: TR/TE = 6312/74 ms, 32-gradient encoding directions b-values = 0 and 1000 s/mm^2^, flip angle = 90°, FOV = 22.4 × 22.4 × 12.0 cm, voxel size= 2×2×2 mm, and 60 slices.

### Supplement C: Linear Mixed Effects Modeling

Linear mixed effects modeling was built in R with the LME4 package. The interaction between the cohort and biological age was tested using the sample code below:

Model 1:
model1 <-lmer(DTI Metric ∼ Age * CHD + Sex + (1|Participant), dataset)
summary(lmer)

Model 2:
model2 <-lmer(DTI Metric ∼ Age + CHD + Sex + (1|Participant), dataset)
summary(lmer)

Likelihood-ratio test:
fm.anova <-anova(model1,model2)
summary(fm.anova)

Predicted Brain Age – Corresponds to the predicted brain age determined by BrainStructuresAges for the structure being analyzed
Cohort – Corresponds to the comparison between cohorts as presented in Table 1.
Sex – Corresponds to the participants’ biological sex
Age – Corresponds to the participants’ chronological age
Participant – Each participant was given a distinct number to account for repeated measures (subject level random intercept).

## Supplementary Results

### Supplement D: Diffusion Tensor Imaging Analysis Supplementary Data

**Figure D1.**
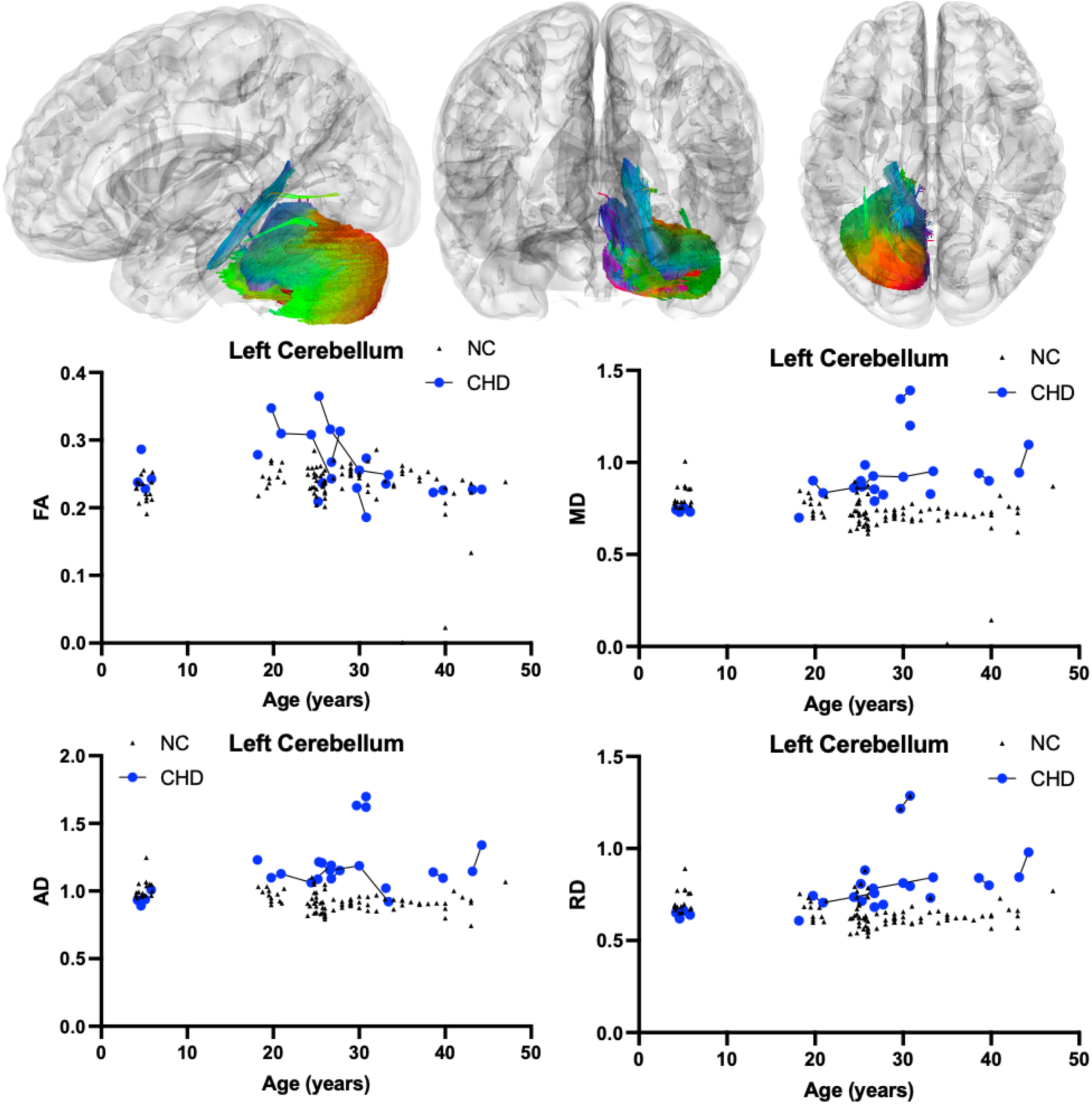
Atlas Based Fiber Tractography of the left cerebellum demonstrating age related effects on A.) fractional anisotropy (FA), B.) mean diffusivity (MD), C.) radial diffusivity (RD), D.) axial diffusivity (AD) between CHD participants (blue) and NC controls (black triangles). There was no statistically significant interaction between participant age and the presence of CHD on left cerebellum FA (*χ*^2^(1) = 0.47, *p*-value_corrected_ = 1.000). There was a statistically significant positive interaction between participant age and the presence of CHD on left cerebellar MD (*χ*^2^(1) = 21.73, *p*-value_corrected_ = 0.0001), AD (*χ*^2^(1) = 15.51, *p*-value_corrected_ = 0.0030), and RD (*χ*^2^(1) = 27.04, *p*-value_corrected_ < 0.0001).

**Figure D2.**
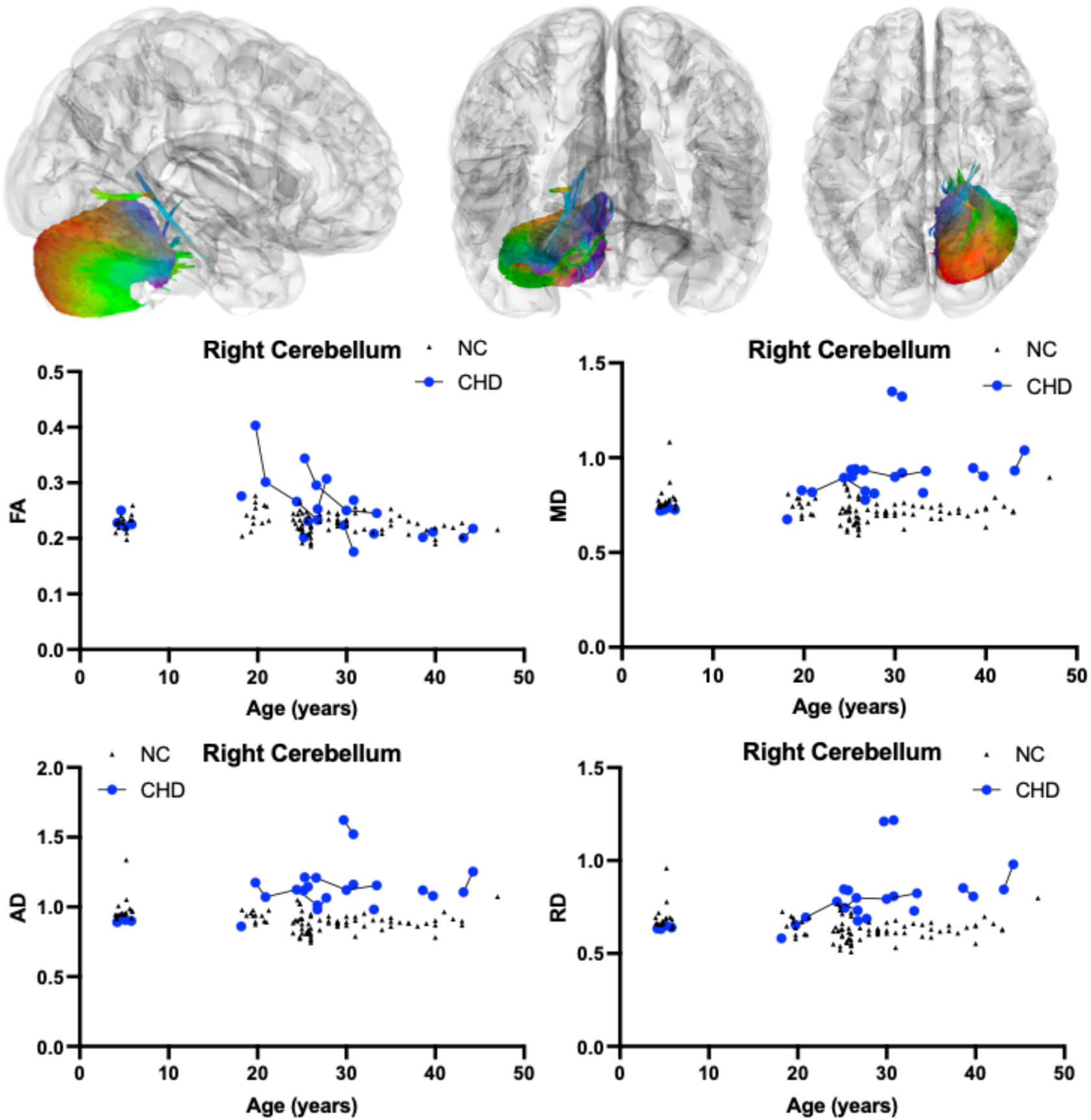
Atlas Based Fiber Tractography of the right cerebellum demonstrating age related effects on A.) fractional anisotropy (FA), B.) mean diffusivity (MD), C.) radial diffusivity (RD), D.) axial diffusivity (AD) between CHD participants (blue) and NC controls (black triangles). There was no statistically significant interaction between participant age and the presence of CHD on right cerebellar FA (*χ*^2^(1) < 0.01, *p*-value_corrected_ = 1.000). There was a statistically significant positive interaction between participant age and the presence of CHD on right cerebellar MD (*χ*^2^(1) = 18.59, *p*-value_corrected_ = 0.0006), AD (*χ*^2^(1) = 13.31, *p*-value_corrected_ = 0.0095), and RD (*χ*^2^(1) = 25.79, *p*-value_corrected_ < 0.0001).

**Figure D3.**
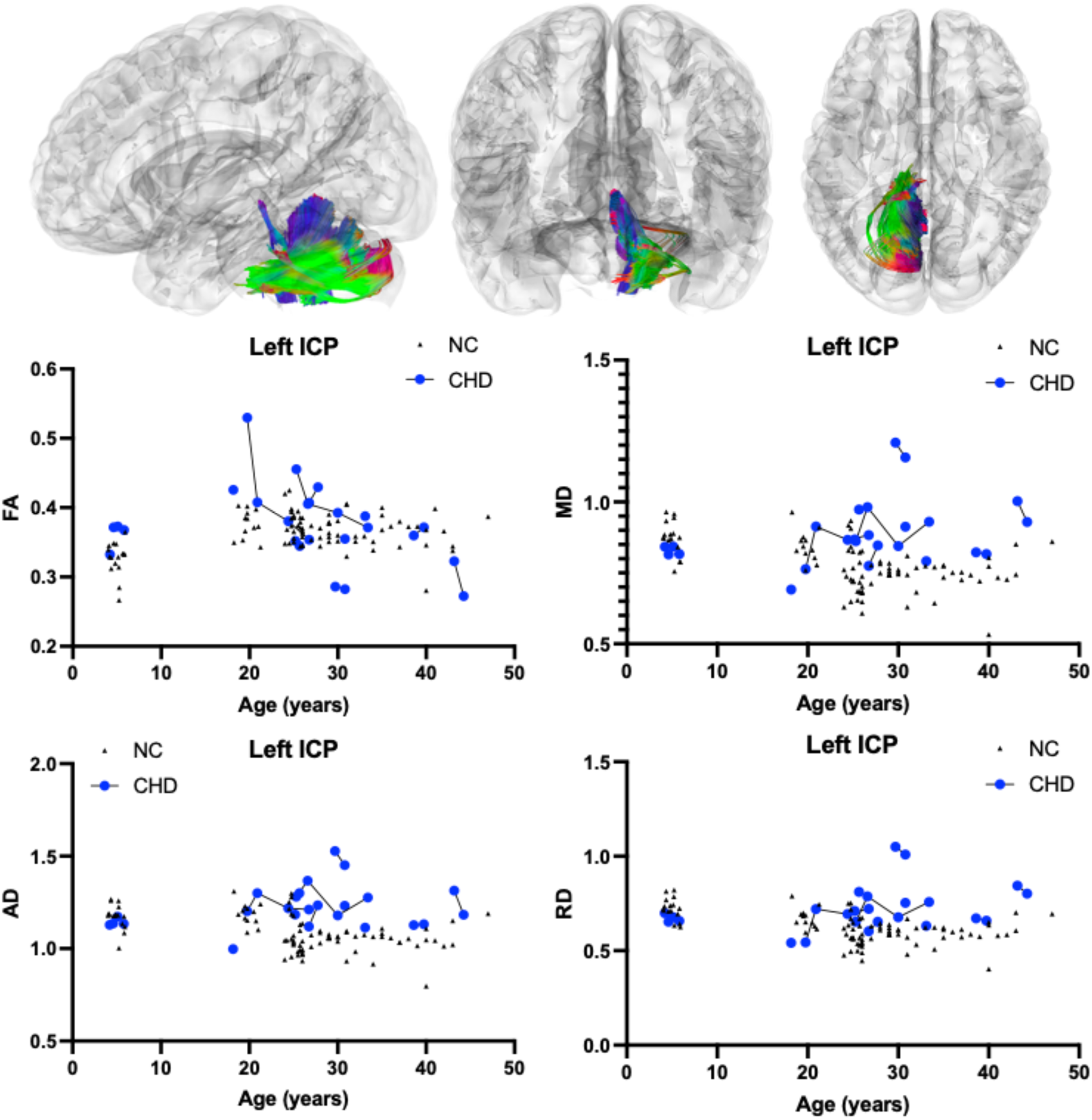
Atlas Based Fiber Tractography of the left inferior cerebellar peduncle (ICP) demonstrating age related effects on A.) fractional anisotropy (FA), B.) mean diffusivity (MD), C.) radial diffusivity (RD), D.) axial diffusivity (AD) between CHD participants (blue) and NC controls (black triangles). There was no statistically significant interaction between participant age and the presence of CHD on left ICP FA (*χ*^2^(1) = 0.79, *p*-value_corrected_ = 1.000) or AD (*χ*^2^(1) = 6.672, *p*-value_corrected_ = 0.3537). There was a statistically significant positive interaction between participant age and the presence of CHD on left ICP MD (*χ*^2^(1) = 13.06, *p*-value_corrected_ = 0.0109) and RD (*χ*^2^(1) = 16.84, *p*-value_corrected_ = 0.0015).

**Figure D4.**
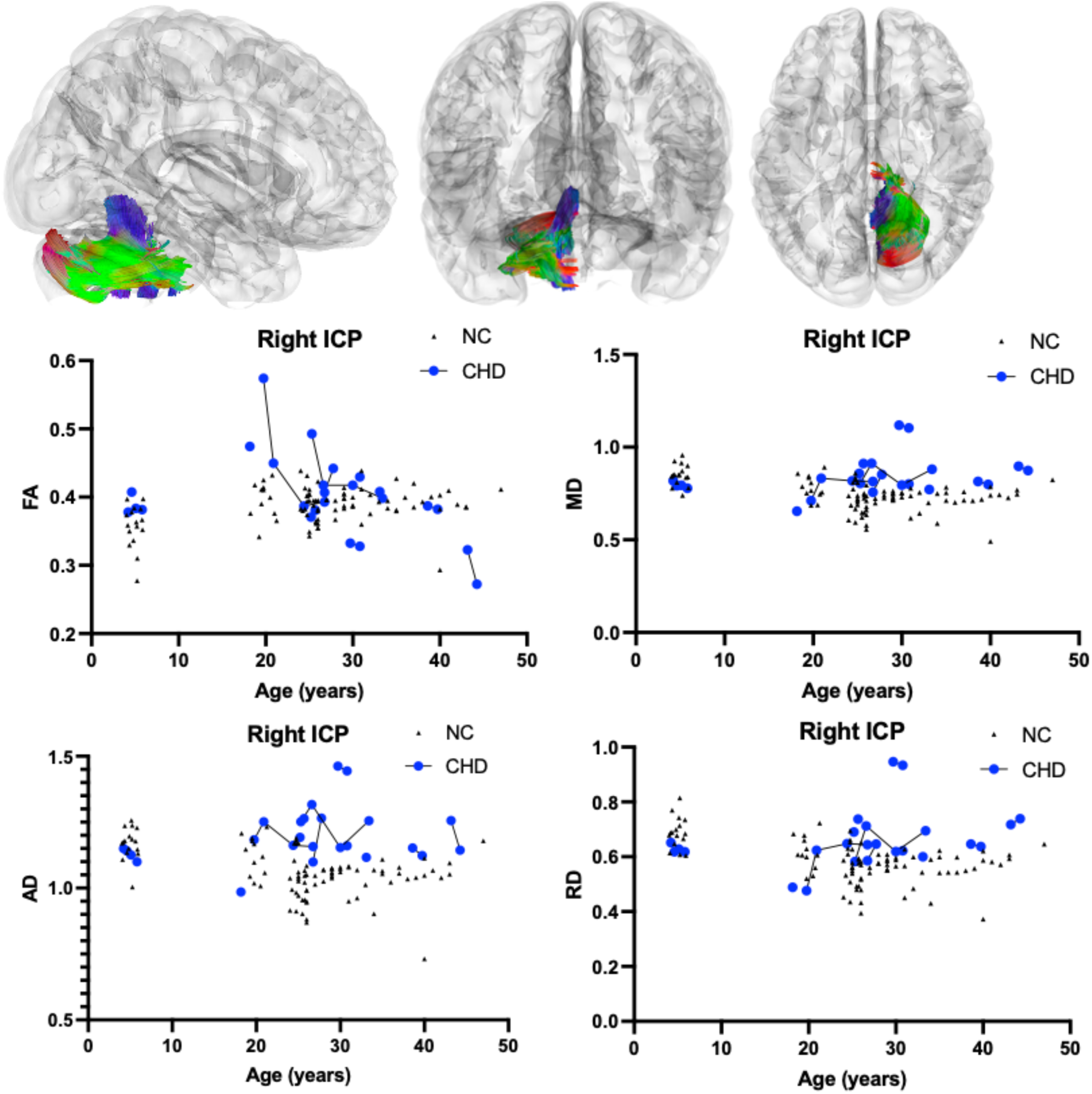
Atlas Based Fiber Tractography of the right inferior cerebellar peduncle (ICP) demonstrating age related effects on A.) fractional anisotropy (FA), B.) mean diffusivity (MD), C.) radial diffusivity (RD), D.) axial diffusivity (AD) between CHD participants (blue) and NC controls (black triangles). There was no statistically significant interaction between participant age and the presence of CHD on right ICP FA (*χ*^2^(1) = 0.30, *p*-value_corrected_ = 1.000) or AD (*χ*^2^(1) = 5.723, *p*-value_corrected_ = 0.6030). There was a statistically significant positive interaction between participant age and the presence of CHD on right ICP MD (*χ*^2^(1) = 12.39, *p*-value_corrected_ = 0.0155) and RD (*χ*^2^(1) = 15.65, *p*-value_corrected_ = 0.0027).

**Figure D5.**
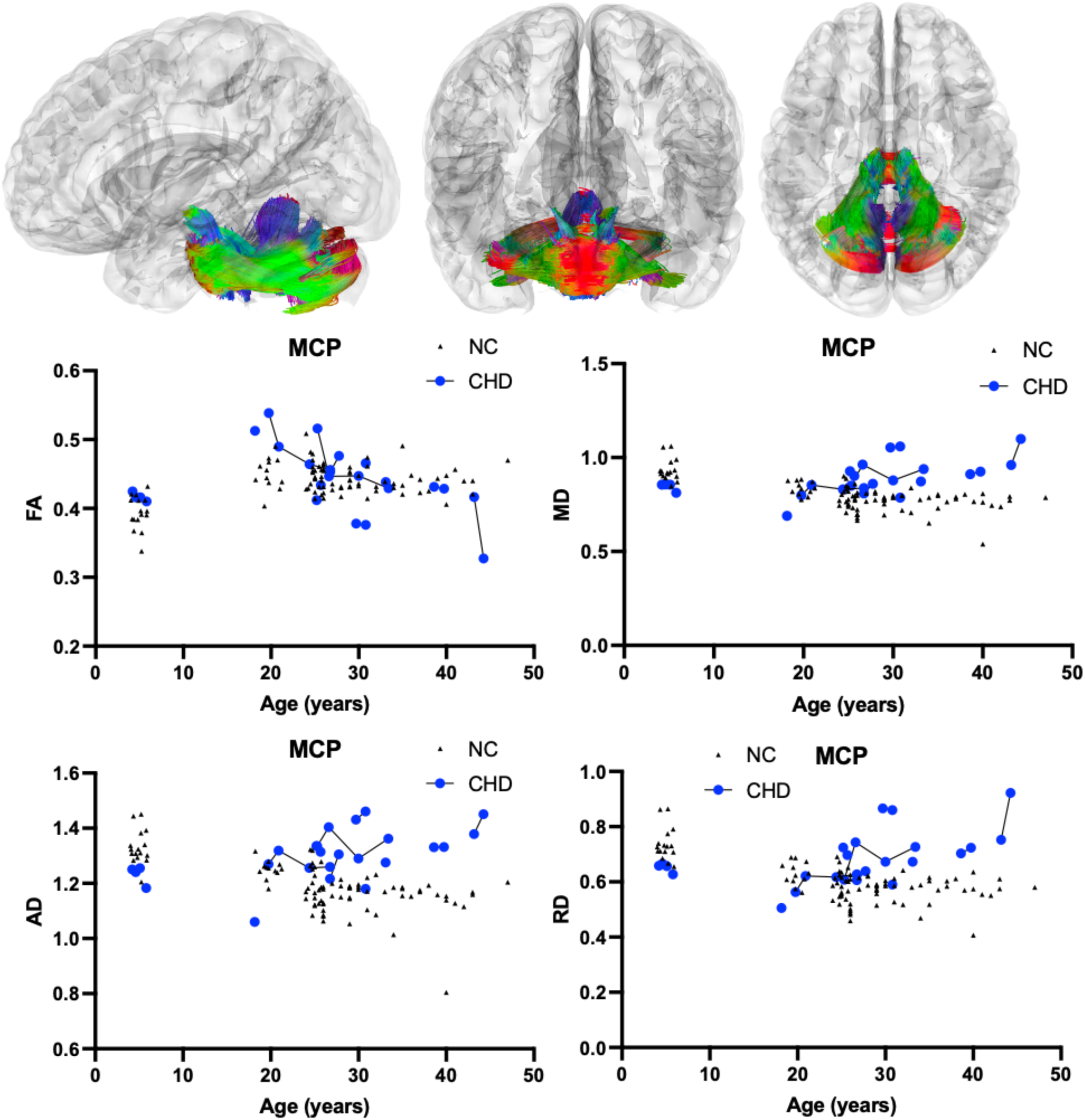
Atlas Based Fiber Tractography of the middle cerebellar peduncle (MCP) demonstrating age related effects on A.) fractional anisotropy (FA), B.) mean diffusivity (MD), C.) radial diffusivity (RD), D.) axial diffusivity (AD) between CHD participants (blue) and NC controls (black triangles). There was no statistically significant interaction between participant age and the presence of CHD on MCP FA (*χ*^2^(1) = 0.31, *p*-value_corrected_ = 1.000). There was a statistically significant positive interaction between participant age and the presence of CHD on MCP MD (*χ*^2^(1) = 20.23, *p*-value_corrected_ = 0.0002), AD (*χ*^2^(1) = 11.66, *p*-value_corrected_ = 0.0230), and RD (*χ*^2^(1) = 24.89, *p*-value_corrected_ < 0.0001).

**Figure D6.**
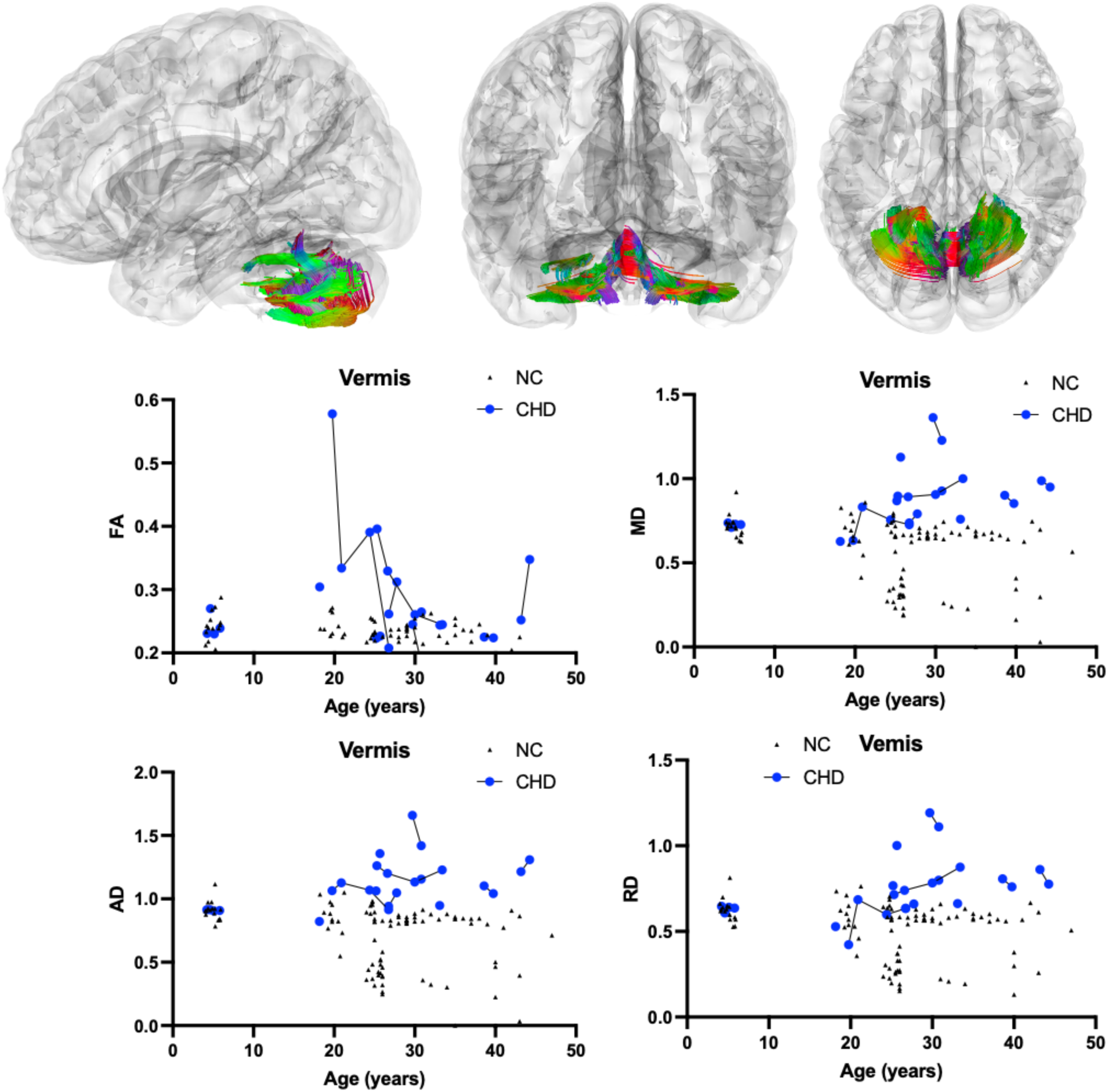
Atlas Based Fiber Tractography of the vermis demonstrating age related effects on A.) fractional anisotropy (FA), B.) mean diffusivity (MD), C.) radial diffusivity (RD), D.) axial diffusivity (AD) between CHD participants (blue) and NC controls (black triangles). There was no statistically significant interaction between participant age and the presence of CHD on vermis FA (*χ*^2^(1) = 1.46, *p*-value_corrected_ = 1.000) or AD (*χ*^2^(1) = 8.783, *p*-value_corrected_ = 0.109). There was a statistically significant positive interaction between participant age and the presence of CHD on vermis MD (*χ*^2^(1) = 13.16, *p*-value_corrected_ = 0.0103) and RD (*χ*^2^(1) = 14.02, *p*-value_corrected_ = 0.0072).

### Supplement E: DTI LMEM Estimates, Standard Errors, and Graphs

**Supplement Table E1.**
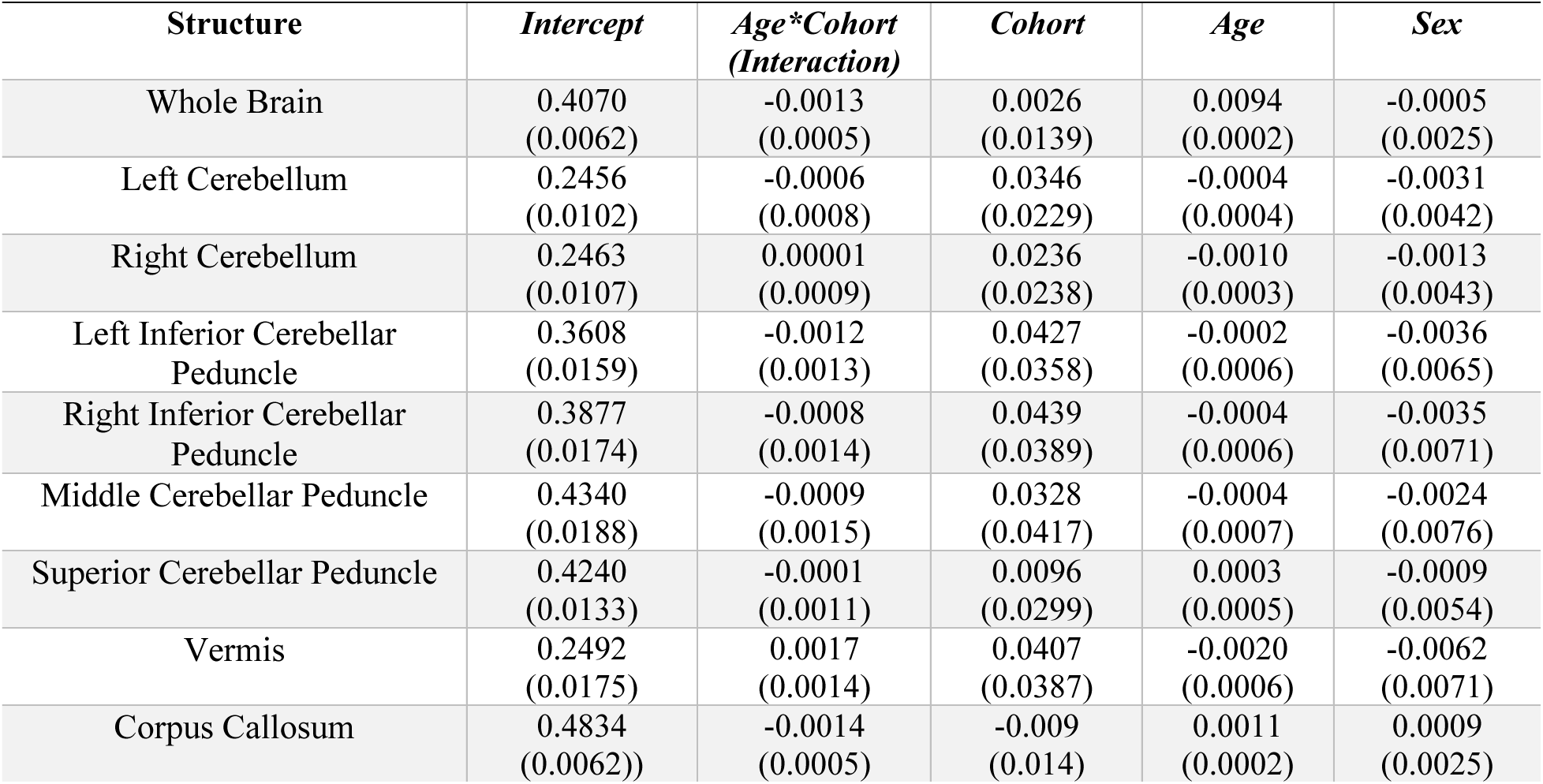
Estimates and Standard Errors from the LMEM for comparisons of FA between CHD and NC for each of the structures analyzed. Estimates are given followed by the standard errors in parenthesis.

| **Structure** | ***Intercept*** | ***Age\*Cohort***<br>***(Interaction)*** | ***Cohort*** | ***Age*** | ***Sex*** |
| --- | --- | --- | --- | --- | --- |
| Whole Brain | 0.4070<br>(0.0062) | -0.0013<br>(0.0005) | 0.0026<br>(0.0139) | 0.0094<br>(0.0002) | -0.0005<br>(0.0025) |
| Left Cerebellum | 0.2456<br>(0.0102) | -0.0006<br>(0.0008) | 0.0346<br>(0.0229) | -0.0004<br>(0.0004) | -0.0031<br>(0.0042) |
| Right Cerebellum | 0.2463<br>(0.0107) | 0.00001<br>(0.0009) | 0.0236<br>(0.0238) | -0.0010<br>(0.0003) | -0.0013<br>(0.0043) |
| Left Inferior Cerebellar<br>Peduncle | 0.3608<br>(0.0159) | -0.0012<br>(0.0013) | 0.0427<br>(0.0358) | -0.0002<br>(0.0006) | -0.0036<br>(0.0065) |
| Right Inferior Cerebellar<br>Peduncle | 0.3877<br>(0.0174) | -0.0008<br>(0.0014) | 0.0439<br>(0.0389) | -0.0004<br>(0.0006) | -0.0035<br>(0.0071) |
| Middle Cerebellar Peduncle | 0.4340<br>(0.0188) | -0.0009<br>(0.0015) | 0.0328<br>(0.0417) | -0.0004<br>(0.0007) | -0.0024<br>(0.0076) |
| Superior Cerebellar Peduncle | 0.4240<br>(0.0133) | -0.0001<br>(0.0011) | 0.0096<br>(0.0299) | 0.0003<br>(0.0005) | -0.0009<br>(0.0054) |
| Vermis | 0.2492<br>(0.0175) | 0.0017<br>(0.0014) | 0.0407<br>(0.0387) | -0.0020<br>(0.0006) | -0.0062<br>(0.0071) |
| Corpus Callosum | 0.4834<br>(0.0062)) | -0.0014<br>(0.0005) | -0.009<br>(0.014) | 0.0011<br>(0.0002) | 0.0009<br>(0.0025) |

**Supplement Table E2.** Estimates and Standard Errors from the LMEM for comparisons of MD between CHD and NC for each of the structures analyzed. Estimates are given followed by the standard errors in parenthesis.

| **Structure** | ***Intercept*** | ***Age\*Cohort***<br>***(Interaction)*** | ***Cohort*** | ***Age*** | ***Sex*** |
| --- | --- | --- | --- | --- | --- |
| Whole Brain | 0.8988<br>(0.0092) | 0.0065<br>(0.0008) | -0.0604<br>(0.0207) | -0.0029<br>(0.0003) | -0.0017<br>(0.0038) |
| Left Cerebellum | 0.8333<br>(0.0294) | 0.0113<br>(0.0024) | -0.1201<br>(0.0641) | -0.0041<br>(0.0011) | -0.0007<br>(0.0120) |
| Right Cerebellum | 0.8144<br>(0.0307) | 0.0105<br>(0.0024) | -0.0959<br>(0.0653) | -0.0040<br>(0.0011) | -0.0033<br>(0.0125) |
| Left Inferior Cerebellar<br>Peduncle | 0.9208<br>(0.0316) | 0.0094<br>(0.0026) | -0.1181<br>(0.0699) | -0.0062<br>(0.0012) | -0.0058<br>(0.0129) |
| Right Inferior Cerebellar<br>Peduncle | 0.8843<br>(0.0315) | 0.0091<br>(0.0026) | -0.1221<br>(0.0691) | -0.0061<br>(0.0012) | -0.0043<br>(0.0129) |
| Middle Cerebellar Peduncle | 0.9794<br>(0.0306) | 0.0114<br>(0.0025) | -0.1879<br>(0.0668) | -0.0076<br>(0.0011) | -0.0036<br>(0.0125) |
| Superior Cerebellar Peduncle | 1.1237<br>(0.0260) | 0.0101<br>(0.0021) | -0.1375<br>(0.0582) | -0.0044<br>(0.0010) | 0.0067<br>(0.0106) |
| Vermis | 0.7539<br>(0.0487) | 0.0141<br>(0.0039) | -0.0598<br>(0.1046) | -0.0067<br>(0.0018) | -0.0199<br>(0.0199) |
| Corpus Callosum | 0.9432<br>(0.0105) | 0.0056<br>(0.0009) | -0.0551<br>(0.0235) | -0.0026<br>(0.0004) | -0.0020<br>(0.0043) |

**Supplement Table E3.** Estimates and Standard Errors from the LMEM for comparisons of AD between CHD and NC for each of the structures analyzed. Estimates are given followed by the standard errors in parenthesis.

| **Structure** | ***Intercept*** | ***Age\*Cohort***<br>***(Interaction)*** | ***Cohort*** | ***Age*** | ***Sex*** |
| --- | --- | --- | --- | --- | --- |
| Whole Brain | 1.3088<br>(0.0106) | 0.0070<br>(0.0009) | -0.0807<br>(0.0235) | -0.0030<br>(0.0004) | -0.0027<br>(0.0043) |
| Left Cerebellum | 1.0432<br>(0.0351) | 0.0119<br>(0.0029) | -0.1009<br>(0.0782) | -0.0049<br>(0.0013) | -0.0036<br>(0.0143) |
| Right Cerebellum | 1.0173<br>(0.0370) | 0.0118<br>(0.0031) | -0.0922<br>(0.0824) | -0.0056<br>(0.0151) | -0.0056<br>(0.0151) |
| Left Inferior Cerebellar<br>Peduncle | 1.2552<br>(0.0421) | 0.0089<br>(0.0034) | -0.0868<br>(0.0926) | -0.0073<br>(0.0015) | -0.0091<br>(0.0172) |
| Right Inferior Cerebellar<br>Peduncle | 1.2282<br>(0.04321) | 0.0084<br>(0.0035) | -0.0716<br>(0.0946) | -0.0073<br>(0.0016) | -0.0076<br>(0.0176) |
| Middle Cerebellar Peduncle | 1.4103<br>(0.0445) | 0.0114<br>(0.0033) | -0.1738<br>(0.0915) | -0.0094<br>(0.0016) | -0.0057<br>(0.0182) |
| Superior Cerebellar Peduncle | 1.6108<br>(0.0369) | 0.0132<br>(0.0031) | -0.1944<br>(0.0830) | -0.0051<br>(0.0014) | 0.0082<br>(0.0150) |
| Vermis | 0.9562<br>(0.0587) | 0.0147<br>(0.0048) | -0.0053<br>(0.1293) | -0.0086<br>(0.0021) | -0.0233<br>(0.0239) |
| Corpus Callosum | 1.4899<br>(0.0101) | 0.0060<br>(0.0008) | -0.0970<br>(0.0223) | -0.0028<br>(0.0004) | -0.0008<br>(0.0041) |

**Supplement Table E4.** Estimates and Standard Errors from the LMEM for comparisons of RD between CHD and NC for each of the structures analyzed. Estimates are given followed by the standard errors in parenthesis.

| **Structure** | ***Intercept*** | ***Age\*Cohort***<br>***(Interaction)*** | ***Cohort*** | ***Age*** | ***Sex*** |
| --- | --- | --- | --- | --- | --- |
| Whole Brain | 0.6937<br>(0.0097) | 0.0064<br>(0.0008) | -0.0514<br>(0.0219) | -0.0029<br>(0.0004) | -0.0013<br>(0.0040) |
| Left Cerebellum | 0.7283<br>(0.0267) | 0.0117<br>(0.0021) | -0.1403<br>(0.0580) | -0.0036<br>(0.0010) | < 0.0001<br>(0.0109) |
| Right Cerebellum | 0.7136<br>(0.0277) | 0.0115<br>(0.0021) | -0.1319<br>(0.0585) | -0.0034<br>(0.0010) | -0.0031<br>(0.0113) |
| Left Inferior Cerebellar<br>Peduncle | 0.7528<br>(0.0271) | 0.0094<br>(0.0022) | -0.1261<br>(0.0604) | -0.0056<br>(0.0010) | -0.0040<br>(0.0110) |
| Right Inferior Cerebellar<br>Peduncle | 0.7121<br>(0.0265) | 0.0089<br>(0.0022) | -0.1334<br>(0.0591) | -0.0055<br>(0.0010) | -0.0027<br>(0.0108) |
| Middle Cerebellar Peduncle | 0.7643<br>(0.0247) | 0.0105<br>(0.0020) | -0.1753<br>(0.0552) | -0.0067<br>(0.0009) | -0.0026<br>(0.0101) |
| Superior Cerebellar Peduncle | 0.8789<br>(0.0222) | 0.0086<br>(0.0018) | -0.1156<br>(0.0494) | -0.0040<br>(0.0008) | 0.0057<br>(0.0090) |
| Vermis | 0.6544<br>(0.0435) | 0.0136<br>(0.0036) | -0.0895<br>(0.0958) | -0.0057<br>(0.0016) | -0.0179<br>(0.0178) |
| Corpus Callosum | 0.6704<br>(0.0120) | 0.0054<br>(0.0010) | -0.0325<br>(0.0270) | -0.0025<br>(0.0004) | -0.0027<br>(0.0049) |

### Supplement F: DTI LMEM Graphs

**Figure F1.**
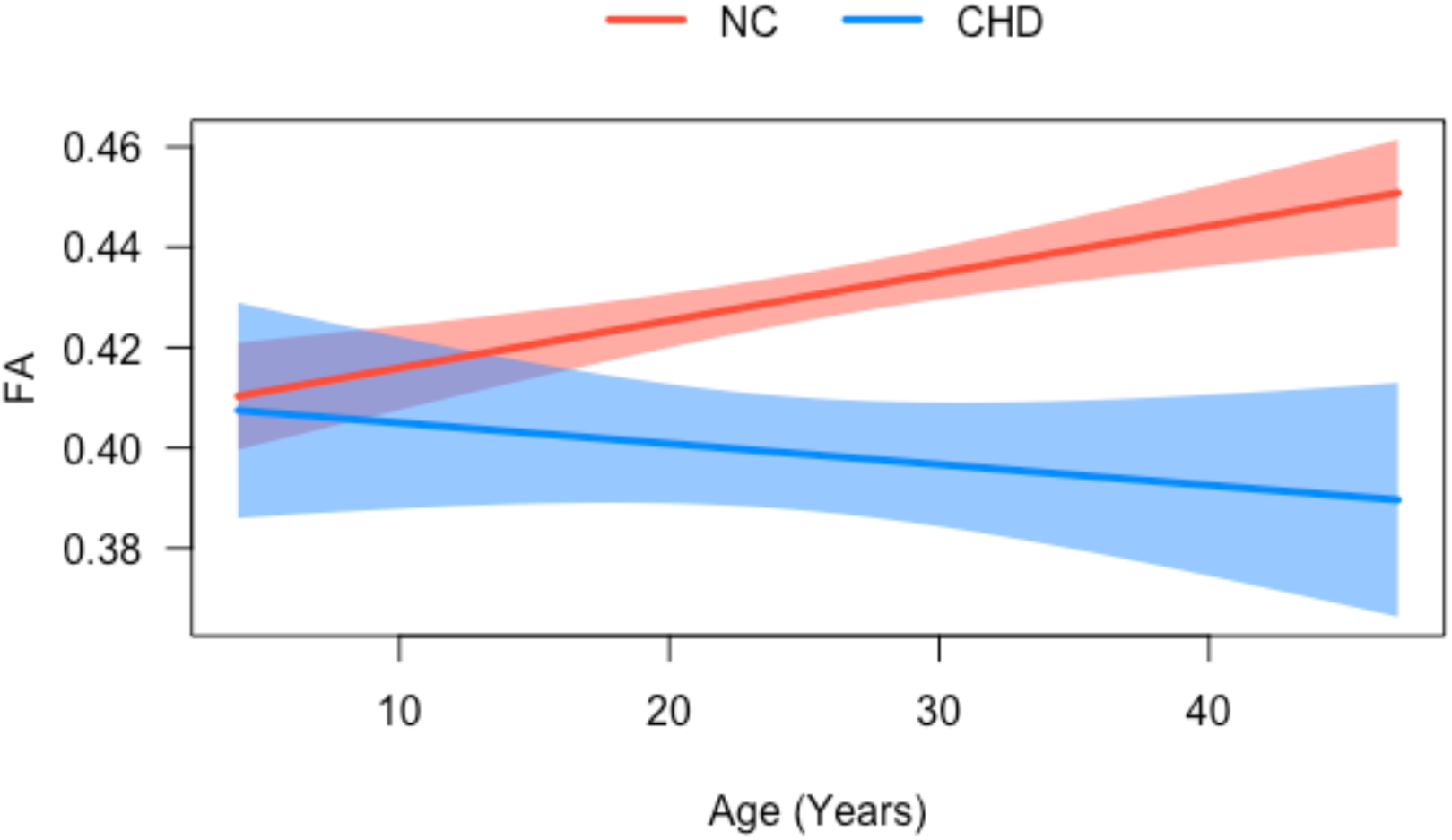
Whole Brain Differences in FA between CHD and NC.

**Figure F2.**
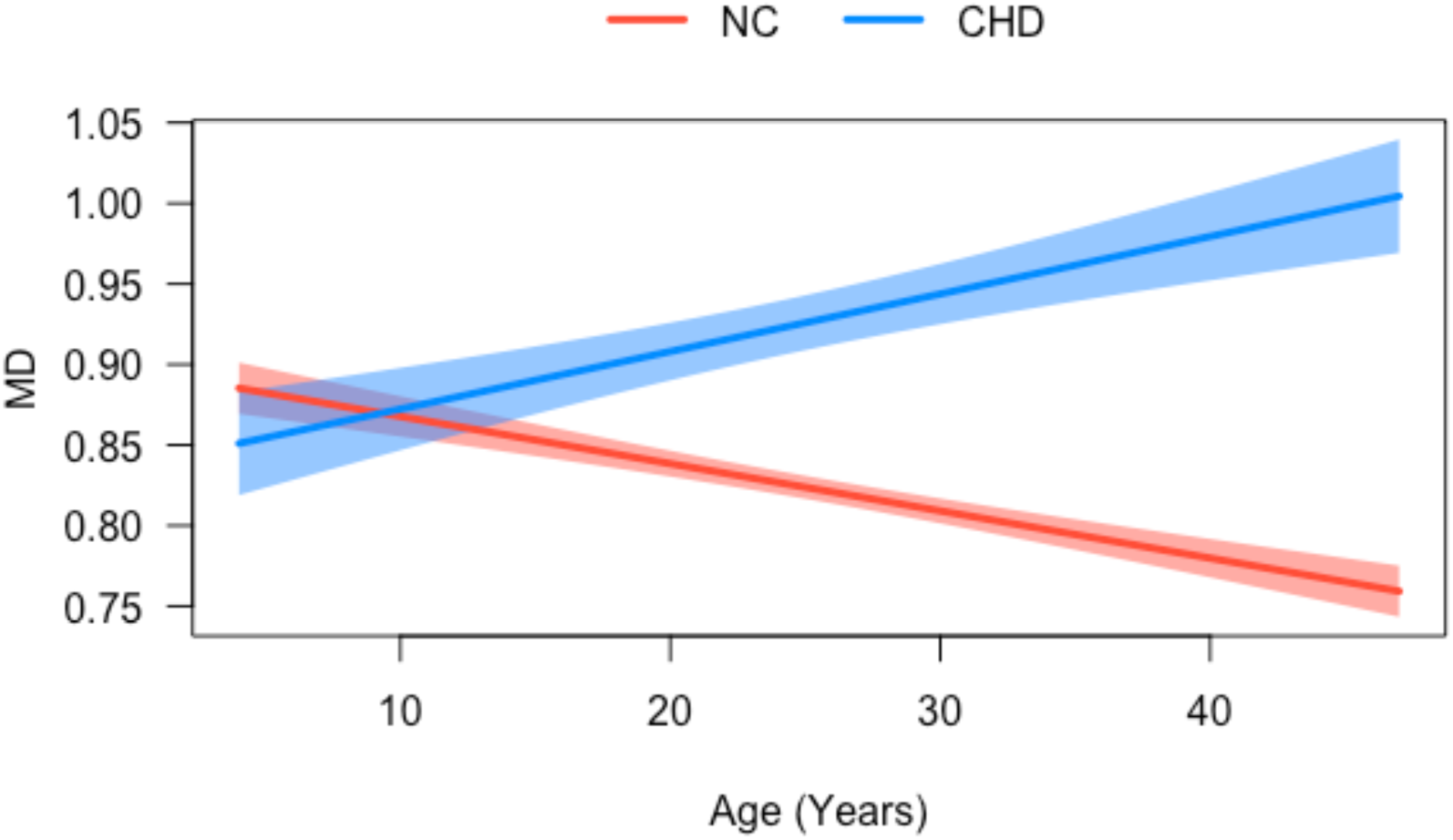
Whole Brain Differences in MD between CHD and NC.

**Figure F3.**
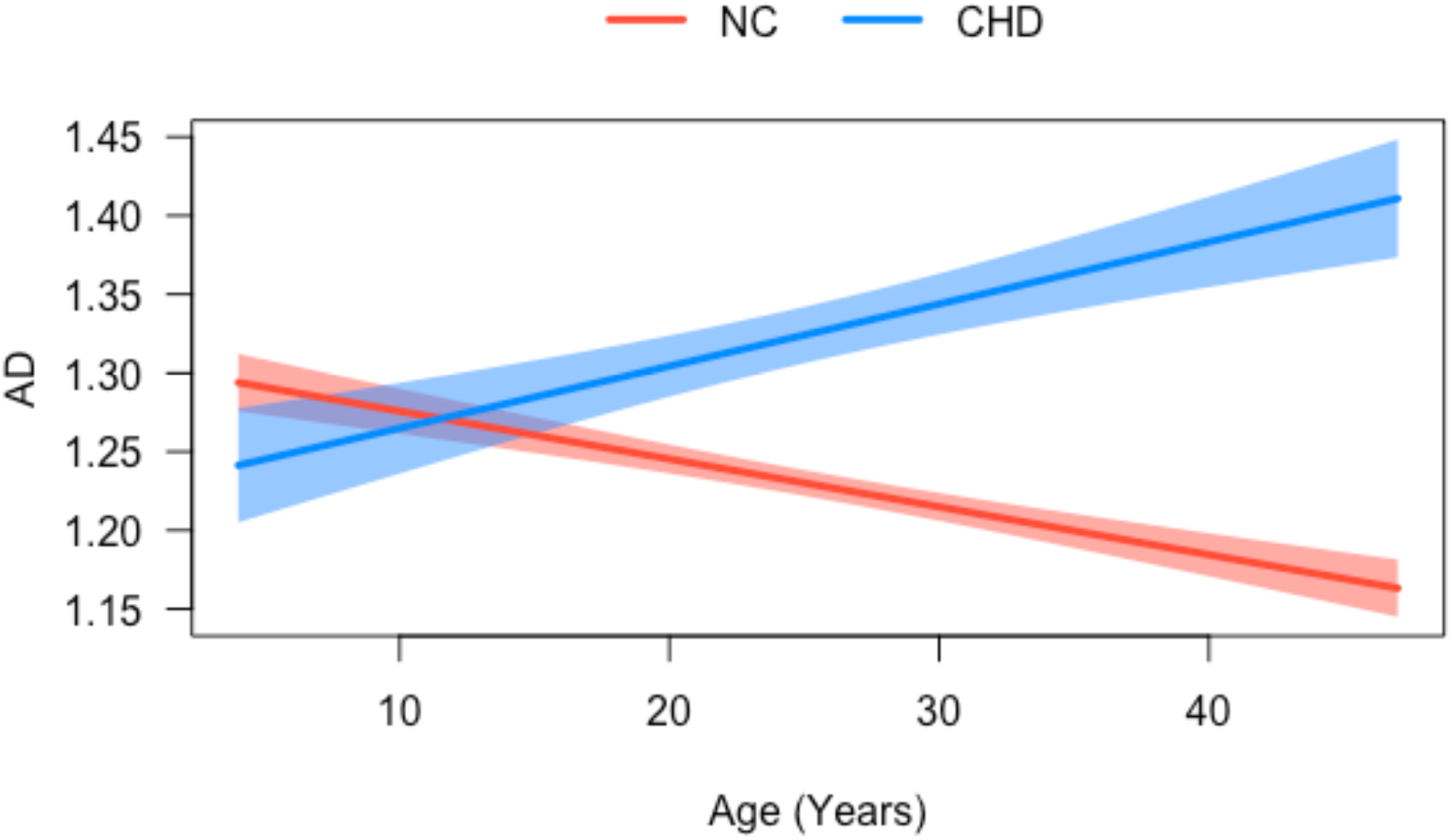
Whole Brain Differences in AD between CHD and NC.

**Figure F4.**
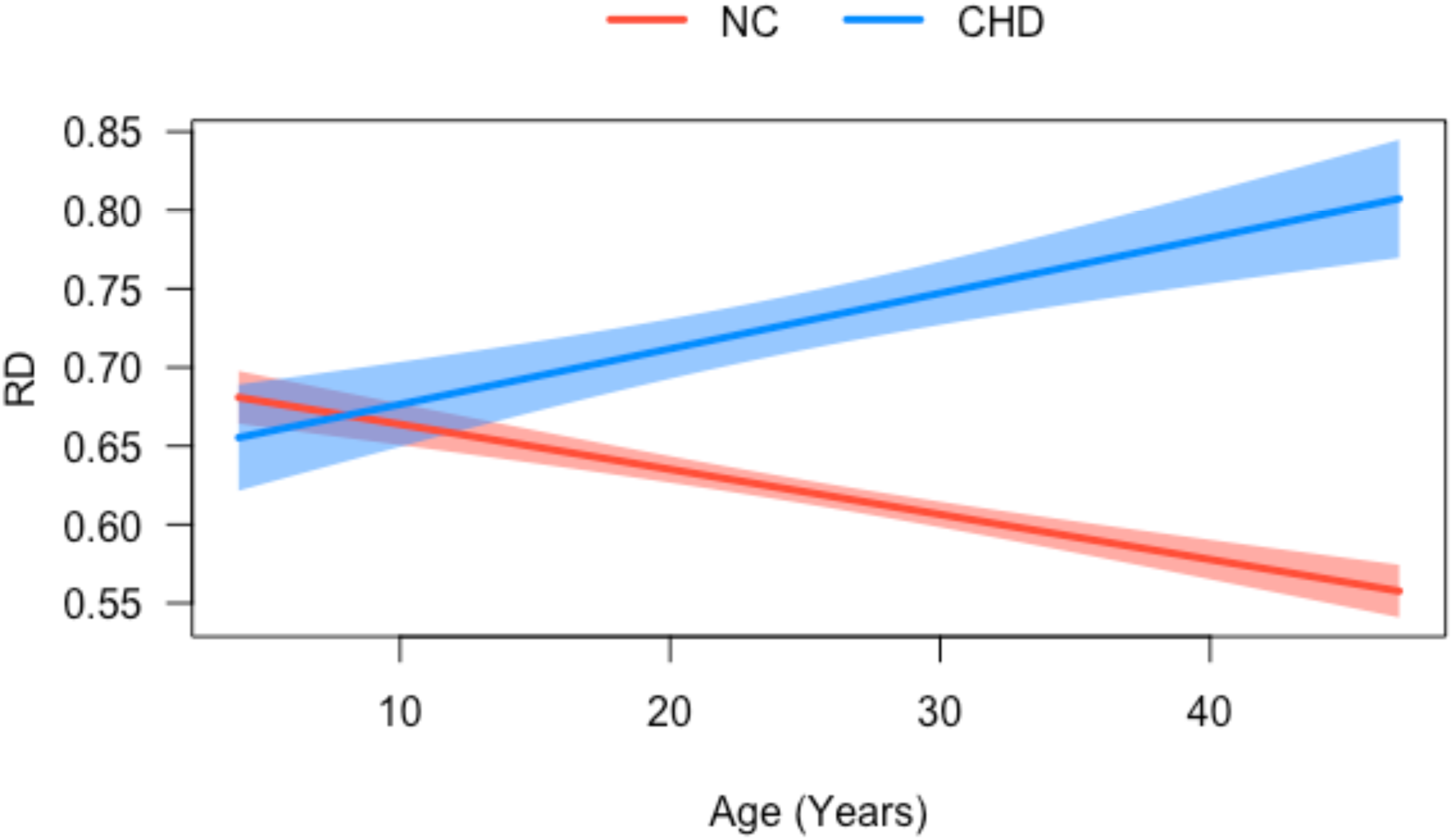
Whole Brain Differences in RD between CHD and NC.

### Supplement G: Correlational Fiber Tractography Supplementary Data

**Figure G1.**
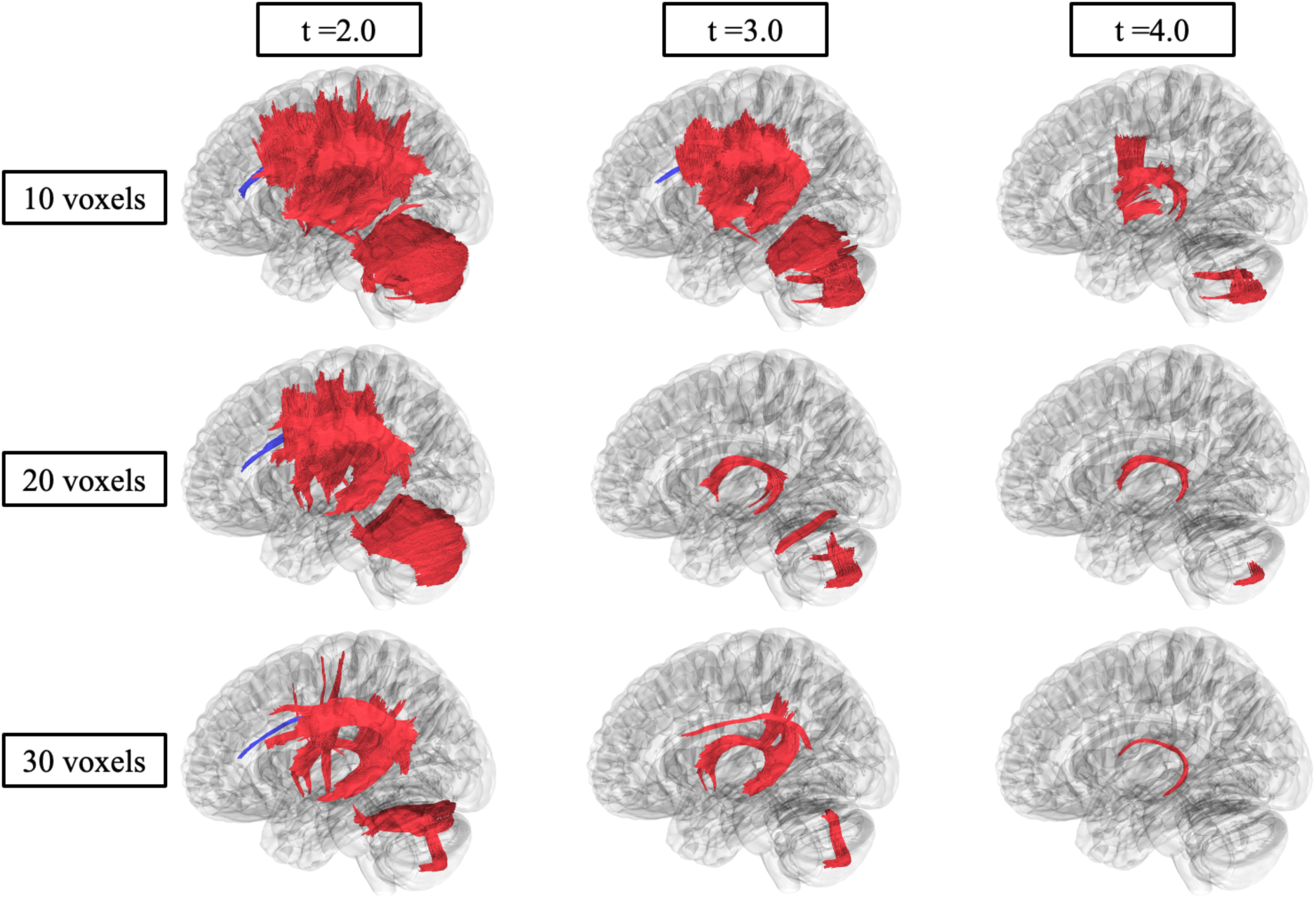
Correlational fiber tractography assessed differences in axial diffusivity (AD) in CHD participants and NC at varying length (voxels) and T threshold. Fiber tracts shown in red were evaluated to have a higher axial diffusivity in CHD participants compared to NC participants and were observed primarily in the cerebellar and corpus callosum pathways (FDR < 0.05). Sparse fiber tracts shown in blue were evaluated to have a higher AD in NC compared to CHD participants and were primarily found in association pathways.

**Figure G2.**
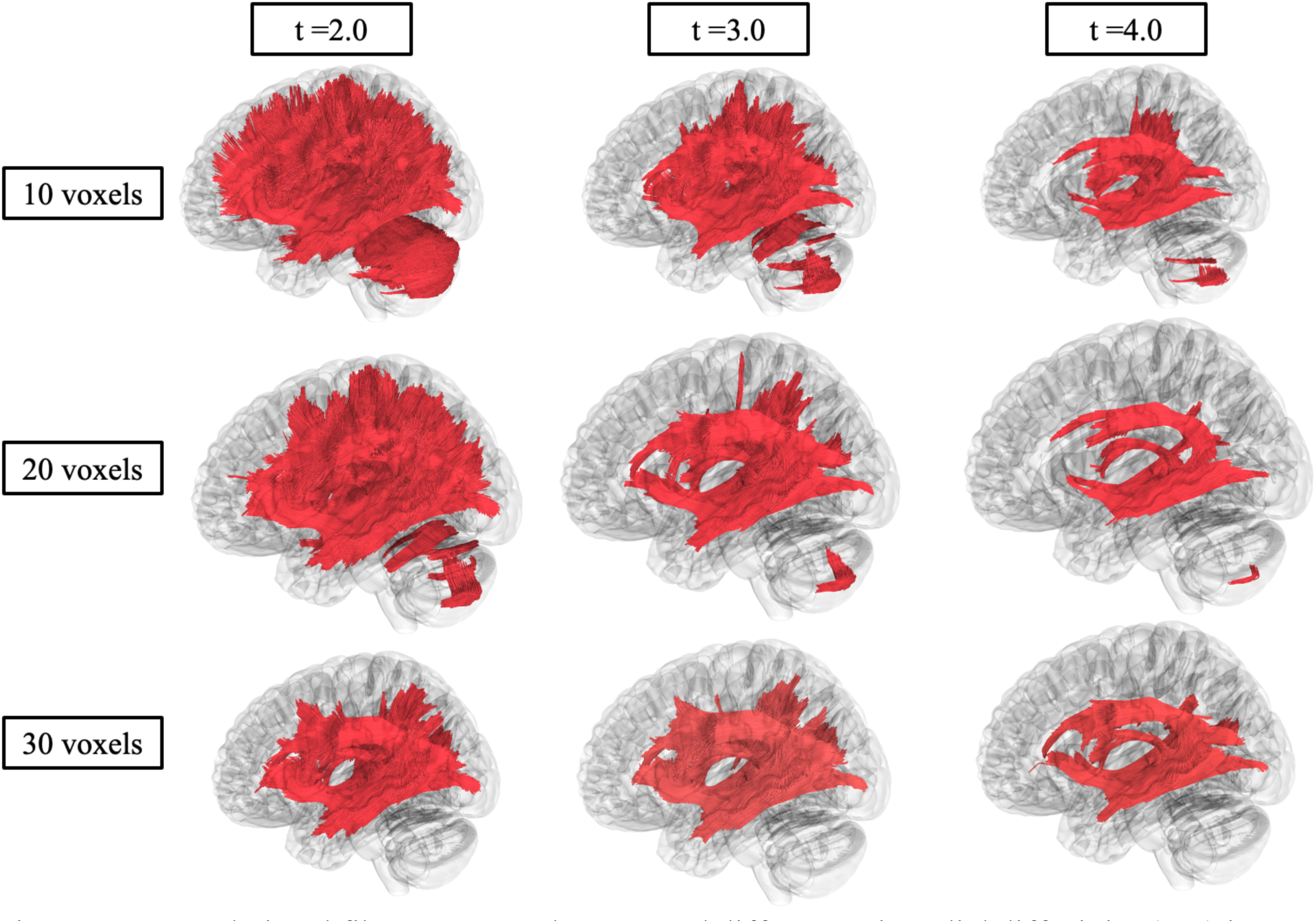
Correlational fiber tractography assessed differences in radial diffusivity (RD) in CHD participants and NC at varying length (voxels) and T threshold. Fiber tracts shown in red were evaluated to have a higher radial diffusivity in CHD participants compared to NC participants and were observed primarily in corpus callosum, cerebellar, and association pathways (FDR < 0.05). No fiber tracts were evaluated to have a higher RD in NC compared to CHD participants.

### Supplement H: Correlational Fiber Tractography Localization

**Table H1:** Fiber Tract Localization for Correlational Fiber Tractography Analysis run with a *T*-score = 2.0 and a length threshold of 10 voxels.

|  | Higher in CHD | Higher in NC |
| --- | --- | --- |
| **FA** | 112845 Tracts<br>Association: 54.06%<br>Commissure: 32.15%<br>Basal Ganglia: 11.09%<br>Brainstem: 2.45%<br>Cranial Nerves: 0.14%<br>Cerebellum: 0.02% | 4932 Tracts<br>Cerebellar: 96.43%<br>Brainstem: 3.57% |
| **MD** | 409376 Tracts<br>Association: 33.18%<br>Commissure: 19.50%<br>Basal Ganglia: 17.28%<br>Brainstem: 15.68%<br>Cerebellar: 14.01%<br>Cranial Nerves: 0.17% | 0 Tracts |
| **AD** | 134693 Tracts<br>Cerebellar: 48.29%<br>Basal Ganglia: 18.61%<br>Brainstem: 17.56%<br>Association: 7.65%<br>Commissure: 7.00%<br>Cranial Nerve: 0.16% | 1663 Tracts<br>100% Association |
| **RD** | 382315 Tracts<br>Association: 42.89%<br>Commissure: 24.42%<br>Basal Ganglia: 15.31%<br>Cerebellar: 8.81%<br>Brainstem: 8.55%<br>Cranial Nerve: 0.17% | 0 Tracts |

**Table H2:** Fiber Tract Localization for Correlational Fiber Tractography Analysis run with a *T*-score = 4.0 and a length threshold of 30 voxels.

|  | Higher in CHD | Higher in NC |
| --- | --- | --- |
| **FA** | 2916 Tracts<br>Association: 92.22%<br>Commissure: 5.08%<br>Basal Ganglia: 2.71% | 0 Tracts |
| **MD** | 31029 Tracts<br>Association: 92.79%<br>Commissure: 1.80%<br>Basal Ganglia: 5.29%<br>Brainstem 0.10% | 0 Tracts |
| **AD** | 87 Tracts<br>Basal Ganglia: 100% | 0 Tracts |
| **RD** | 30865 Tracts<br>Association: 82.90%<br>Basal Ganglia: 10.31%<br>Commissure: 6.77%<br>Brainstem: 0.01% | 0 Tracts |

## Notes

### Competing Interest Statement

The authors have declared no competing interest.

### Clinical Protocols

https://clinicaltrials.gov/study/NCT00005917

### Author Declarations

The National Institutes of Health Institutional Review Board approved this protocol (00-HG-0153). Written informed consent was obtained from patients or the legal guardians of the patients. Informed consent was completed before participation and all research was completed in accordance with the Declaration of Helsinki.

